# Prospective study on the organization and efficiency of online journal club

**DOI:** 10.64898/2026.08.11.26360192

**Authors:** Nikita N. Burlov, Matvei Y. Baranovskii, Elizaveta A. Burlova, Matvei G. Slavenko, Gleb N. Khrykov

## Abstract

**Background:** Journal clubs (JCs) are a popular education format. Interest in studying their impact is high, and authors often report positive results related to subjective parameters. Objective assessments of effectiveness are limited and contradictory. In this paper, we share our experience and describe our journal club’s effectiveness.

**Methods:** We conducted a prospective cohort study within our online journal club. Meetings followed a discussion-based format and were held via Zoom, with timing and topics determined by voting in the club’s Telegram chat. Enrolment occurred in waves and included an application, entry test, and interview. During each recruitment wave, both club members (treatment group) and applicants (control) completed an admission test assessing knowledge of evidence-based medicine and statistics.

**Results:** The JC currently comprises 27 members. Over the past year, 76 meetings were held, with 75% of participants grading their experience with 9 or 10 on a ten-point scale. Multivariate analysis demonstrated non-significantly results (SMD = 0.19 (95% CI 0.004; 0.38), p = 0.046) among participants. However, in other adjusted models, differences between groups were not statistically significant (p > 0.05).

**Conclusion:** While the analysis of the subjective outcomes is consistent with findings from previous studies, the objective outcomes remain inconclusive. Further research is needed to refine the methodology for the organization and evaluation of journal clubs.

## Background

A journal club (JC) is a group of medical professionals meeting regularly to discuss articles of interest from medical journals [1]. These meetings are typically devoted to the critical reading of research papers, aimed at enhancing participants’ understanding of study design, statistical methods, and critical assessment skills [2]. Generally, JCs are not part of the compulsory medical curriculum; rather, they provide an additional opportunity for knowledge maintenance, staying updated with recent research, and developing skills in methodology and data analysis. Both organizing and participating in a JC require a considerable investment of time [1]. Some authors describe their experience of hosting JCs in the form of structured courses for university students or residents [3–6]. However, evaluating JCs as independent educational interventions remains challenging, partially due to heterogeneity in their organization and stated objectives being too vague (e.g. “critical reading” or “changing medical practice”). Consequently, no consensus has been reached regarding the optimal format of JCs or the most effective methods for assessing their impact.

Some studies have reported improvements associated with journal club participation, including increased reading time (from 2.0 hours to 3.5 hours, p = 0.026) as well as higher knowledge and critical appraisal scores (from 50.8 to 62.9, p = 0.003) [7]. In 2008, Deenadayalan Y. et al. published a meta-analysis reporting increase in knowledge acquisition and critical reading skills [2]. However, other studies found no statistically significant differences in knowledge levels of the journal club participants: MS1 cohort (mean (SD)): pre-test 12.24 (1.71), post-test 11.41 (2.15) p = 0.187; MS2 cohort: pre-test 13.17 (1.95), post-test 13.33 (1.87) p = 0.949 [5]. Similarly, another study reports: pre-intervention: TREAT group M = 63.8, Standard group M = 66.2 post-intervention: TREAT group M = 68.1, Standard group M = 66.9. p = 0.167 [8]. A recent meta-analysis further supports these findings (Std. Mean Difference = 0.15 (95% CI −0,09; 0,39), p = 0.22) [9].

Currently, there are few publications on the experience of organizing journal clubs in Russia. These publications do not report on effectiveness assessments, only on participant characteristics [10-12]. Nonetheless, JCs are popular and widely practiced in the country, both as extracurricular activities within universities and independently outside academia. To the best of our knowledge, the most closely related study was published in 2021 by K. Baron and colleagues from the Higher School of Oncology, a Russian training center for oncology residents. The study demonstrated that online teaching by foreign experts improved participants’ knowledge of evidence-based medicine and patient communication. However, since the JC was but one of several educational methods, the effectiveness of the JC itself could not be assessed [13].

This paper aims to present our experience in organizing and conducting our JC, as well as assessing its effectiveness.

## Material and methods

When organizing our JC, we employed the theoretical framework proposed by M. Linzer et al., which posits that a general medicine JC can affect clinical practice by promoting the habit of reading research articles, enhancing knowledge in epidemiology and biostatistics, and developing critical appraisal skills [14]. The JC was launched in October 2020, and data collection commenced at that time, with the study designed prospectively prior to the first meeting.

### The Structure of the Journal Club

The JC meetings are conducted online, providing more convenience for participants and enabling broader reach. Eligibility for admission is extended to medical students in their fourth (out of six) year or later (corresponding to the start of clinical training in the university curriculum), residents, and practicing physicians. Besides that, professionals from fields related to medicine, such as veterinaries, medical journalists, biologists and medical educator are also eligible to apply.

Admission to the JC is conducted in waves, i.e. defined recruitment periods. Admission requires completing a three-step selection process: (1) submission of an application form, (2) successful completion of tests in evidence-based medicine and statistics (the specific test varies by admission wave) as well as an English language assessment (EF SET 15 Min Quick Check), and (3) an interview with the JC organizers (BN and BE). In the application form (administered via Google Forms), candidates provide basic information (age, profession, place of study or employment, number of publications, prior experience with JCs, and self-assessment of knowledge in evidence-based medicine) and submit a short personal essay. Candidates scoring below 30% on the knowledge tests are not advanced to the interview stage.

The interview is used to clarify the candidate’s goals and motivation, confirm their ability to attend meetings according to the JC schedule, and ensure willingness to adhere to the JC’s rules. Final admission decisions are made collectively by the organizers. No alternative ways of joining the JC are available; all participants may withdraw from the JC whenever they are inclined to do so.

Using the experience from previous research, we outlined a set of rules intended to enhance the effectiveness of the JC [8, 15, 16]:

1. Participants address each other informally (using the second-person singular).
2. The date, time and topic of each meeting are selected by voting in the JC’s Telegram chat (meetings are typically held twice per month).
3. The article for discussion is circulated vie the JC chat at least four days in advance, allowing participants sufficient time to review it.
4. Each meeting is recorded, and both the recording and supplementary materials are made available to members in a shared Google Drive folder.
5. The meeting format, invited speakers, and home task assignments are determined by the organizers, although JC members are encourage to submit their suggestions.
6. Participants are expected to notify the organizers in advance if they anticipate being absent.
7. All questions – regardless of how trivial they may seem – are welcomed and actively encouraged.
8. Attendance and participation (i.e., active contributions during meetings and voting) are monitored. Three consecutive absences result in expulsion, and consistent non-participation despite presence prompts review by the organizers.
9. Meetings emphasize open and unstructured discussion.
10. A designated leader with relevant expertise guides the session.
11. Mentorship, peer teaching, and mutual learning are integral components of the JC.

### Meetings format

All meetings are conducted online via the Zoom client (Zoom Video Communications, Inc.). Most commonly, sessions are organized in a discussion format, in which the supervisor prompts the conversation by posing questions about the article under analysis, and participants respond while critically examining the importance, benefits and disadvantages of each feature of the article. Occasionally, a report format is used, whereby a designated participant presents the main details of the study and subsequently invites the participants to discuss some of its elements. Besides that, the organizers, members, or invited guests (e.g., experienced clinicians domain experts) may give lectures on topics such as evidence-based medicine, research methodology, statistics or any specific areas of medicine (e.g., patient communication, decentralized science etc.). After each meeting, participants may submit an anonymous review evaluating both their overall impression and the perceived usefulness of the knowledge gained, using a Likert scale (ranging from 1 to 10 or from 1 to 5). Participants may also provide open-ended feedback, commenting on aspects they appreciated as well as areas they found unsatisfactory or in need of improvement. In addition, the organizer (BN) occasionally assigns homework to consolidate received knowledge and to promote further learning. Such tasks may include studying a definition, solving theoretical problems, critically evaluating the strengths and weaknesses of an article from a certain perspective, etc.

### Outcomes

The JC has no pre-specified end date; rather, it functions as a community of individuals sharing interests, dedicated to gaining new and maintaining existing knowledge. Hence it is not striving to achieve a single, specific goal. We nonetheless assume and hope that within the aforementioned format the JC is still capable of influencing the participants’ clinical practice [14].

This outcome, however, is difficult to measure or quantify due to its broad definition, and many things come into play, such as Hawthorne effect. A more practical approach might be to examine outcomes at another level, specifically changes in participants’ reading habits and the “effectiveness” of these practices. This, however, assumes either collecting self-reported evidence, which lack objectivity, or on extensive monitoring of participants, which is not feasible due to the online format as well as related ethical concerns. For these reasons, we selected performance on evidence-based medicine and statistics tests as our primary outcome measure, considering these results as indirect surrogates measuring the knowledge gained during the JC membership.

Starting from the second wave of recruitment, all candidates were required to complete a test, and their results were used as the control group. All candidates who correctly submitted the admission form and met the professional eligibility criteria were admitted to the test, hence no additional selection by the organizers occurred. It was prospectively planned that current JC members would also complete the same test. Thus 6 sets of measurements (corresponding to the recruitment waves from the 2^nd^ to 7^th^) were obtained, each comprising two groups: the treatment group (JC members) and the control group (candidates).

To minimize recall bias, different tests were administered during each recruitment wave, and the test names were not disclosed to participants. The following tests were used: 2^nd^ wave (02.2021) – a complex test, including USMLE Sample Test Questions Step 1, 2020; Fresno Test of Evidence Based Medicine; and Coursera platform: Design and Interpretation of Clinical Trials course (maximum score of 11 points); 3^rd^ wave (08.2021) – modified SPLIT (mSPLIT) test with the maximum score of 15 points; 4^th^ wave (02.2022) – ACE tool; 5^th^ wave (11.2022) – Berlin test; 6^th^ wave (10.2023) – RESET test, 7^th^ wave (11.2024) – SPLIT test; 8^th^ wave (02.2026) – ACE tool (full text in Supplementary 1) [17–23]. In addition, the authors did not provide answers for tests in publications. For example, in the SPLIT test, we had to complete the assessment ourselves and consider as correct those answers that were agreed upon by consensus among the researchers. English proficiency was determined according to results of the EF SET test (15 MIN QUICK CHECK, EL test): beginner 1–60%, intermediate 61–85%, advanced 86–100% [24].

### Statistical Analysis

Summary statistics for continuous variables are presented as means (SD) or medians (IQR), and categorical variables are reported as counts and relative frequencies.

In a consensus meeting BN, BM and BE identified potential confounders for the association between recruitment wave and tests results and developed a directed acyclic graph (DAG, see Figure 1). The primary regressor of interest was the membership status in our JC, with candidates serving as controls, and JC members as treated units. The identified covariates included age (continuous), profession (classified as students, residents, postgraduate students, practicing physicians, and other), and previous JC experience (yes / no).

**Figure 1.**
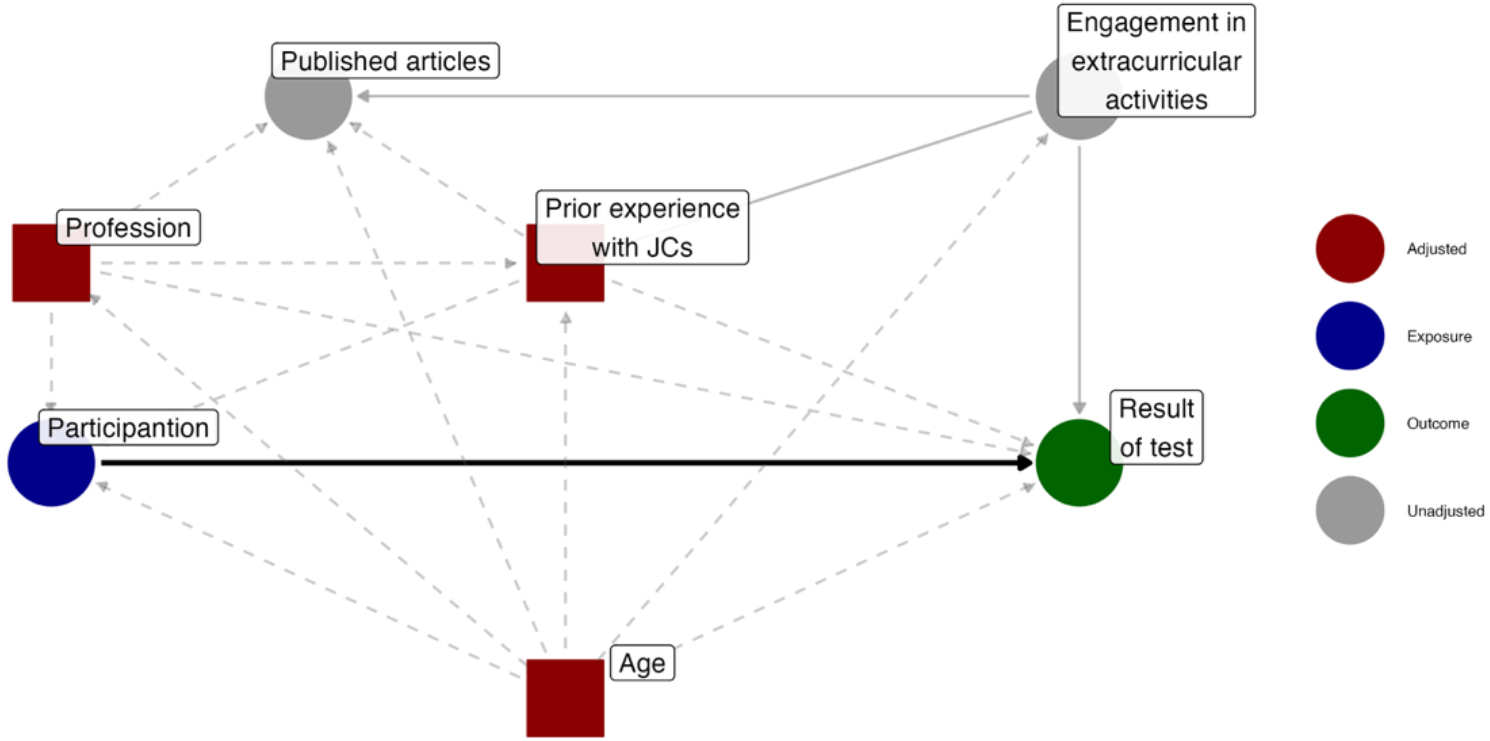
Directed acyclic graph (DAG) for statistical analysis.

A series of uni- and multivariable linear regressions were estimated to examine the associations between recruitment wave and RESET test results: univariate Model 1 included only the JC membership status; Model 2 adjusted for age; Model 3 for age and previous experience with JCs; and Model 4 for age, prior JC experience, and profession. Comparing these models allowed to see how the effect estimates and the corresponding uncertainties change. Subsequently, all test results were standardized (z-scores). For each test a linear model was fitted with z-score as response, and JC membership, as well as age, prior JC experience, and profession as regressors (Models 5–11). For Models 1–11, Wald-type 95% confidence intervals (CI) were computed using heteroscedasticity-consistent robust standard errors (type HC3 sandwich estimators) [25].

An additional linear mixed model (Model 12) was fitted, using z-scores from all tests as the response. Fixed effects included age, prior JC experience, profession at the moment of testing (e.g. a student enrolled to the JC may graduate and become a practicing doctor as the time passes), test type, JC membership status, and the interaction between test type and membership status; a random intercept was allowed for each person ID. Standard errors were estimated using cluster-robust variance-covariance estimator with bias-reduced linearization (CR2 type) [26]. Marginal and conditional (given the test type) effects are reported as standardized mean difference (SMD) with 95% CI, derived using a counterfactual grid.

Results were considered statistically significant when the two-sided p-value was lower than 0.05. Statistical analysis was performed using R version 4.5.0 for Mac OS (RStudio IDE version 2025.05.0+496) using the following packages: *readxl, tidyverse, dagitty, ggdag, lme4, sandwich, clubSandwich, gtsummary, modelsummary, marginaleffects, ggdist, ggthemes, patchwork*.

## Results

### General characteristics

The flowchart (Figure 2) illustrates the history of JC and its recruitment process. As of February 2026, there were 24 active members in the JC: 1 (4.2%) from the 1^st^ recruitment wave, 2 (8.3%) from the 4^th^, 1 (4.2%) from the 5^th^, 6 (25%) from the 6^th^, 14 (58%) from the 7^th^. Recruitment wave occurred as follows: November 2020 (1st), March 2021 (2nd), October 2021 (3rd), April 2022 (4th), January 2023 (5th), October 2023 (6th), November 2024 (7th) and February 2026 (8th). During the five years, a total of 105 participants were excluded from the JC due to repeated absences from meetings.

**Figure 2.**
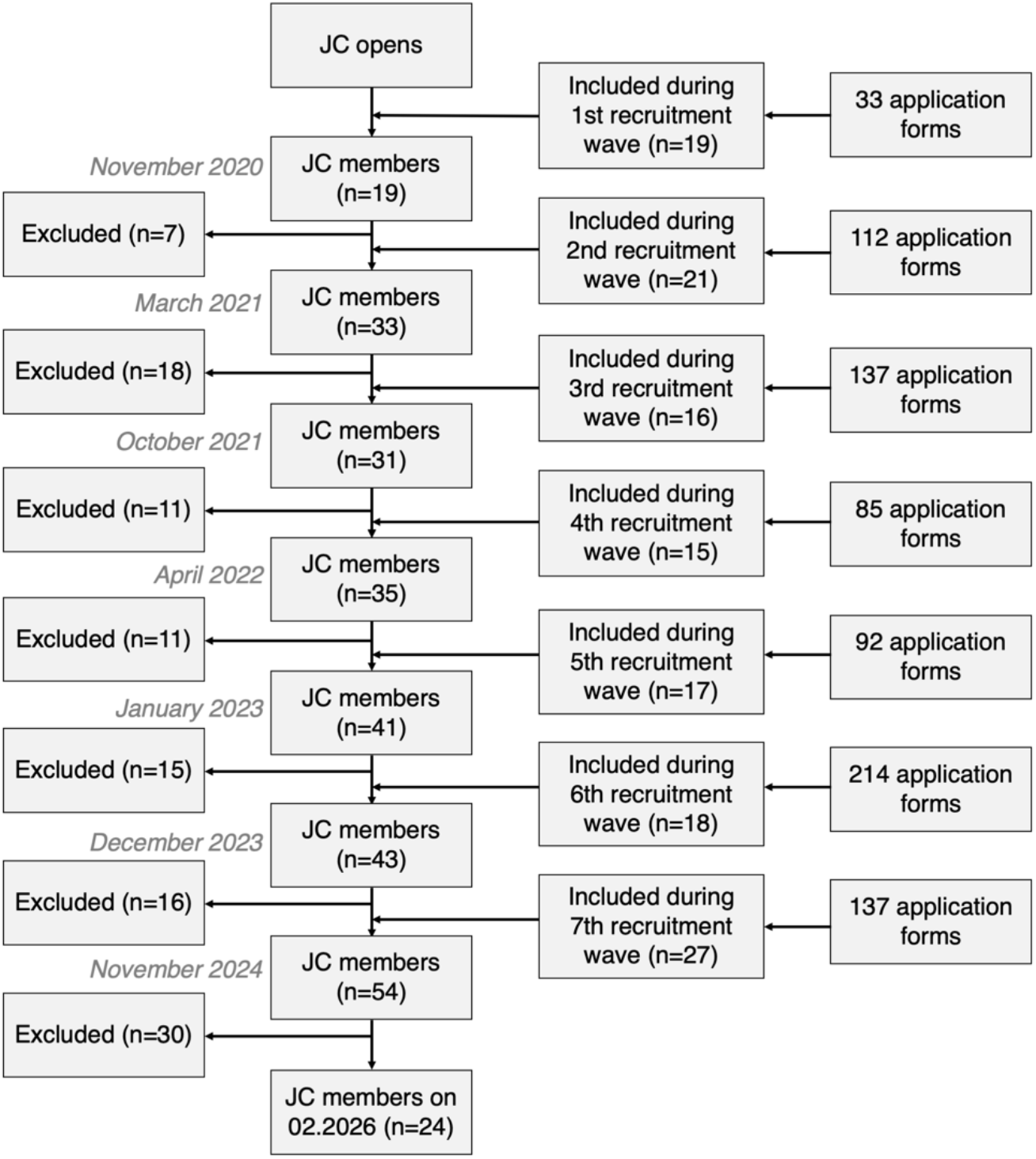
Flow-chart of history journal club.

Table 1 presents the main characteristics of the JC members. Most of them were practicing physicians (n=13, 54%). The mean (SD) age of members was 29.5 (4.8) years. At the moment of enrollment, only 11 (46%) participants had published their own scientific works, primarily in Russian journals. English proficiency was predominantly classified as *Intermediate* (n=10, 42%) and *Advanced* (n=13, 54%).

**Table 1.** Comparison of basic characteristics between “participants” and “non-participants”.

| Characteristic | participants,<br>n = 24 | non-participants,<br>n = 65 |
| --- | --- | --- |
| Age<sup>1</sup> | 29.5 (4.8) | 29.1 (6.1) |
| Sex |  |  |
| Female | 13 (54%) | 45 (69%) |
| **Male** | 11 (46%) | 20 (31%) |
| **Profession** |  |  |
| **Student** | 1 (4.2%) | 9 (14%) |
| **Resident** | 4 (17%) | 18 (28%) |
| **Practicing physicians** | 13 (54%) | 24 (37%) |
| **Postgraduate student** | 2 (8.3%) | 5 (7.7%) |
| **Other** | 4 (17%) | 9 (14%) |
| **Prior experience with JCs** | 12 (50%) | 25 (38%) |
| **Engagement in extracurricular activities** | 15 (63%) | 37 (57%) |
| **Have published articles** | 11 (46%) | 42 (65%) |
| **Number of publications<sup>2</sup>** | 4 (2, 15) | 5 (3, 9) |
| **Have published in international journals** | 2 (8.3%) | 23 (35%) |
| **Knowledge of EBM and statistics before JC** | 22 (92%) | 55 (85%) |
| **English proficiency level** |  |  |
| **Beginner** | 1 (4.2%) | 14 (22%) |
| **Intermediate** | 10 (42%) | 46 (71%) |
| **Advanced** | 13 (54%) | 5 (7.7%) |

<sup>1</sup>Mean (SD); <sup>2</sup>Median (IQR). Other – medical teacher, medical specialist and research assistant.

From November 2020 to February 2026, a total of 96 meetings were held. Of these, 63 (83%) followed a discussion format, 40 (42%) included lectures, and in 4 (4%) meetings a designated participant presented an article (categories not mutually exclusive). Participants of the JC led 18 (19%) meetings, 14 (15%) meetings were held by invited guests, and the rest were led by the organizers. The mean (SD) duration was 131 (22.9) min, with a mean (SD) attendance of 20.7 (7.5) participants per session (median attendance rate: 67%, IQR 57–74%). Participation dynamics is illustrated in Figure 3, and detailed descriptions of each session are available in Supplementary 2.

**Figure 3.**
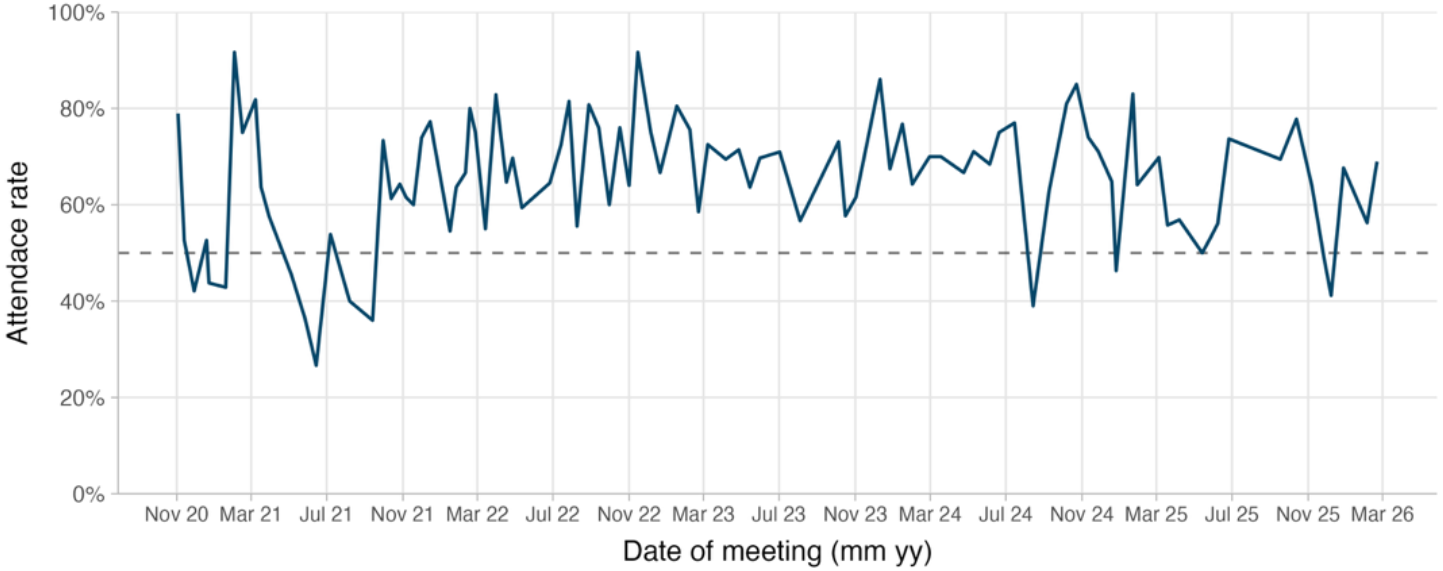
Dynamics of visits meeting.

Dashed line – 50% attendance rate.

Table 1 also summarizes the basic characteristics of the candidates during the 8^th^ recruitment wave. Differences are observed in profession (practicing physicians: 54% vs. 37%, residents: 17% vs. 28%; students: 4.2% vs. 14%), prior JC experience (50% vs. 38%) and English proficiency level.

### Subjective data

We received a total of 425 feedback reviews from the JC members. Participants were very satisfied with the meetings: on a scale from 1 (negative) to 10 (positive) the mean (SD) score was 9.2 (1.0), with 75% of reviews rated at 9 or 10. As for the quality of material presentation, 96% reviews described it as either “optimal” or “simple”.

### Statistical Inference

According to the univariable linear model (Model 1) for the ACE tool, JC members performed non-significantly results than candidates (p = 0.117). After adjusting for the potential confounders (Models 2–4), the difference continued statistically non-significant (Model 4: MD = 0.49 (95% CI -0.28; 1.26), p = 0.210), see Table 2. In the linear mixed model (Model 12), the overall effect of JC participation was statistically, but not clinically significant (SMD = 0.19 (95% CI 0.004; 0.38), p = 0.046); the CI is rather wide, covering both practically negligible effects from 0.004 to 0.1 SDs, as well as potentially substantial shifts of up to 0.38 SDs.

**Table 2.**
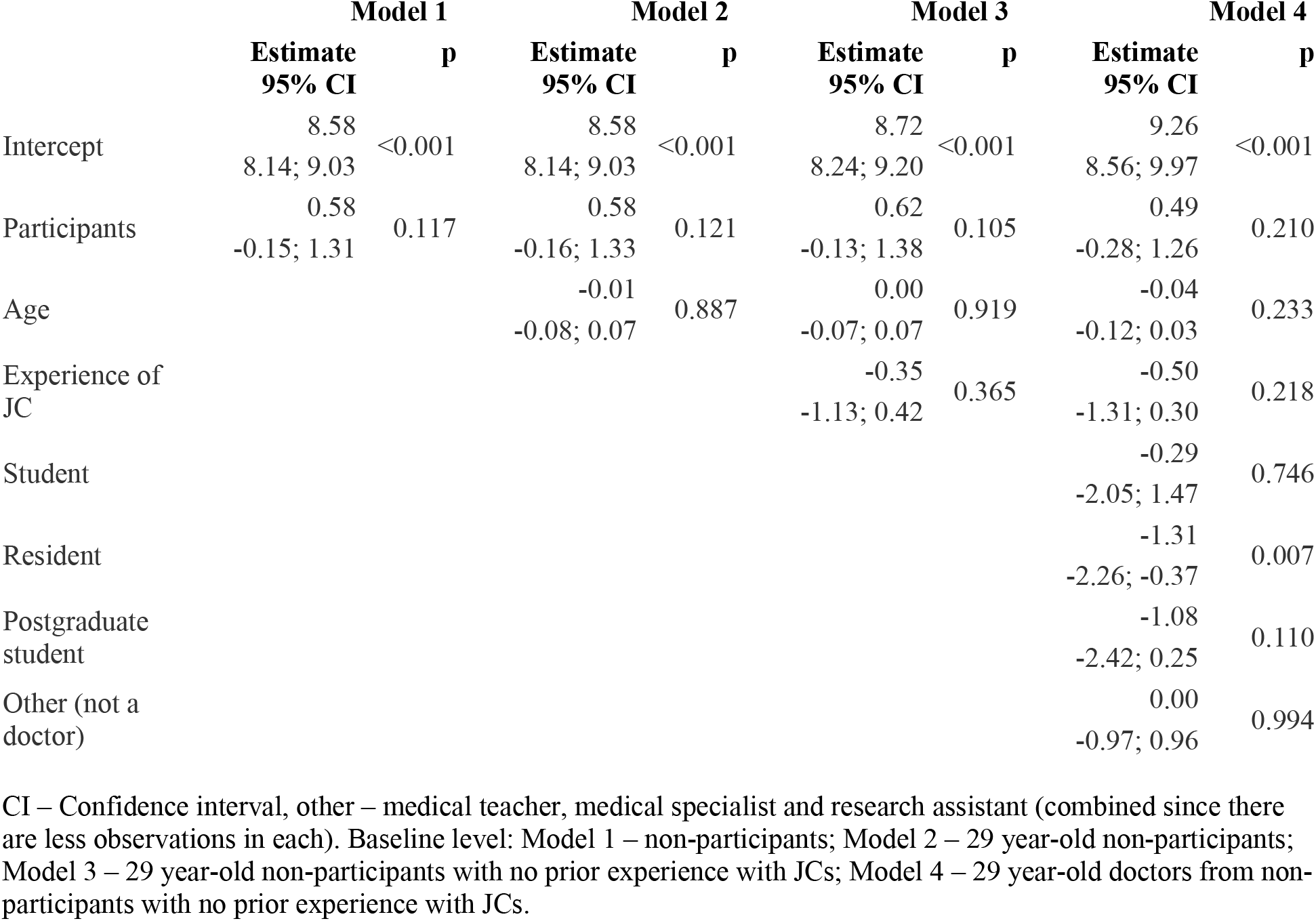
Regression modelling results.

When comparing Models 5–11 and Model 12 (see Table 3 and Figure 4), the following results are of interest. In the Modified SPLIT and ACE tests, the point estimates indicate that, on average, candidates performed better than the JC members, although the difference is not statistically significant. In contrast, for the RESET and SPLIT tests, the JC members outperformed candidates, with the difference reaching statistical significance (RESET: SMD = 0.56, (95% CI 0.22; 0.91), p = 0.001, SPLIT: SMD = 0.49, (95% CI 0.11; 0.87), p = 0.012); nonetheless the CI is too wide for the effect to be practically interpretable.

**Table 3.** Comparison of results produced by simple linear models (Models 5–11) and linear mixed model (Model 12).

| Test | LM (Models 5–11) |  | LMM (Model 12) |  |
| --- | --- | --- | --- | --- |
|  | SMD<br>95% CI | p | SMD<br>95% CI | p |
| **Complex** | 0.35<br>–0.3; 1.01 | 0.287 | 0.43<br>–0.08; 0.94 | 0.098 |
| **Modified SPLIT** | –0.16<br>–0.76; 0.43 | 0.587 | –0.42<br>–0.9; 0.06 | 0.084 |
| **ACE** | –0.27<br>–0.97; 0.43 | 0.448 | –0.44<br>–1.03; 0.14 | 0.138 |
| **Berlin** | 0.13<br>–0.35; 0.6 | 0.604 | 0.19<br>–0.22; 0.6 | 0.356 |
| **RESET** | 0.43<br>–0.05; 0.9 | 0.077 | 0.56<br>0.22; 0.91 | 0.001 |
| **SPLIT** | 0.71<br>0.22; 1.2 | 0.005 | 0.49<br>0.11; 0.87 | 0.012 |
| **ACE** | 0.29<br>–0.16; 0.74 | 0.206 | 0.09<br>–0.34; 0.52 | 0.669 |

SMD – standardized mean difference, CI – confidence interval, LM – linear model, LMM – linear mixed model. Average marginal effects for models 5-11, conditional average marginal effects for model 12.

**Figure 4.**
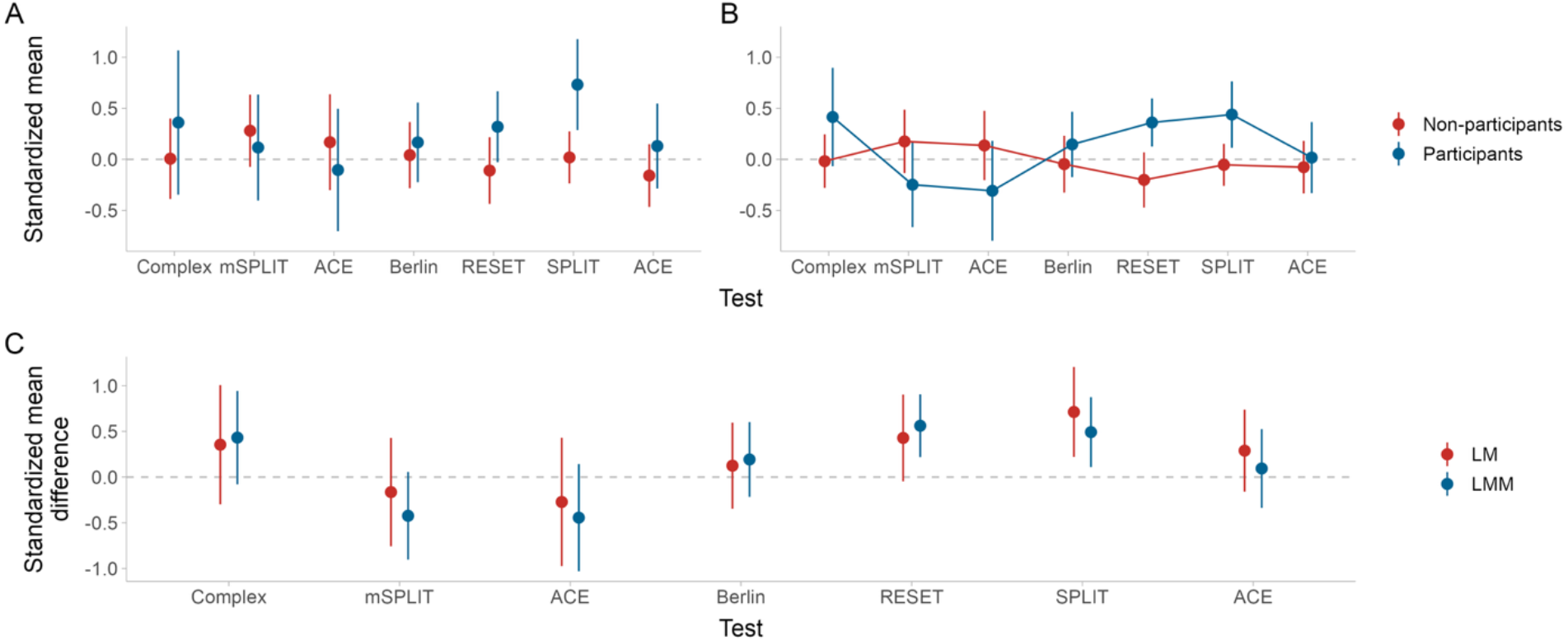
Results of tests. Point – estimates, range – 95% confidence intervals, LM – linear model (Models 5–11), LMM – linear mixed model (Model 12), mSPLIT – Modified SPLIT test. A) Mean standardized test scores for (non-)participants estimated using Models 5–11. B) Mean standardized test score for (non-)particpants estimated using linear mixed model (Model 12). C) Estimated mean difference in standardized test scores between participants and non-participants (Models 5–11).

## Discussion

This study presents an example of organizing an online journal club and provides an assessment of its effectiveness. Although there are many journal clubs in the Russian Federation, both online and in-person, to the best of our knowledge, only the Higher School of Oncology has previously reported their results [13].

In its modern form, the journal club format is usually attributed to William Osler, who initially conceived is as a means of accessing journals he could not afford personally. The first documented meeting titled “The Journal Club” was held on 29 October 1889 in the library of Johns Hopkins Hospital, Baltimore. Back then, JCs were dedicated primarily to studying new publications [27]. Compared to the 19^th^ century, the information nowadays is more readily available, thus the majority of JCs, including ours, mainly put emphasis on cultivating the skill of critical analysis of articles, rather than on providing access to newly published papers.

A deliberate choice in favor of the closed format in our JC was made when it was conceived. It was assumed that requiring candidates to undergo an admission process would enhance the perceived value of membership, thus uniting and motivating the participants. To the best of our knowledge, such an organizational model has never been described in the literature before.

Another distinctive feature of our JC was the breadth of the eligibility criteria. In most articles, the following distinct cohorts are identified: students, residents, practicing physicians [7, 8, 28–30]. Our JC admitted a more heterogeneous group: students from the 4^th^ year onward, residents, physicians, and professionals from fields related to medicine, such as veterinaries, medical journalists, biologists, and medical educators. Despite this diversity, residents and physicians constituted the majority of members, reflecting both their greater interest and their higher levels of engagement. Notably, most of them joined the JC as students or residents, and their professional status evolved during their membership. This heterogeneity has its advantages: students and residents get an opportunity to interact with practicing physicians, ask questions and learn how the methods from articles are applied in practice. Physicians, in turn, get a chance to practice their teaching and mentoring skills, and support junior colleagues in the beginning of the career path.

At the moment of writing, our JC comprised 27 participants. The largest number of members reported in the literature was described by Wenke et al., who documented 126 clinicians. This study however combined the experiences of nine JCs [8]. Most published articles describe groups of 20–30 participants, a range comparable to the size of our JC [11, 31, 32].

The scheduling of meetings – frequency, date and time – was aimed at maximizing the attendance. In accordance with the recommendations by Aronson, our meetings were held regularly (2–3 times a month), although without a fixed time [16]. We believe this flexible yet frequent format leads to a more effective acquisition of knowledge and practice of the applied skills. This view is confirmed by self-reports and voting of our members. Nonetheless, other studies suggest that a fixed date contributes to the JC effectiveness [8, 15]. In our case, the average attendance rate was approximately 67%, which exceeded the threshold of 50% we deemed necessary for sustaining the JC. Although we did not formally compare attendance with a control group, prior studies report higher attendance rates in JCs with more structured organization [14, 28]. However, these studies are relatively dated, which may limit the generalizability of their findings to modern JCs.

To assess the effectiveness of participation in our JC, we used scores from tests on EBM and statistics. The tests were adapted to the background knowledge of our participants and applicants, since some tests were, in our opinion, excessively complex and advanced for our target audience. We observed statistically significant differences between participants and applicants in the sixth seventh recruitment waves; however, in eighth wave these differences not preserved in univariable model and after adjusting on potential confounders. In the mixed-effects linear model with adjustment on confounders, participants performed statistically significantly better in RESET and SPLIT test. This may suggest that long-term participation in a JC contributes positively to knowledge acquisition. Nevertheless, the corresponding CI is wide, which precludes firm conclusions regarding the practical significance of the effect; perhaps the further research will provide more evidence in favour of this hypothesis.

We offer several explanations for the obtained results: 1) our modifications of the tests may have compromised their validity as measures of knowledge in statistics and EBM and, thereby, limiting their utility as indicators of effectiveness; and 2) each new recruitment wave may have attracted applicants who were progressively more interested and better prepared in these subjects.

Only a limited number of studies report objective assessments of participants’ knowledge, and the available findings remain inconsistent [7, 8, 14, 21–23, 33]. To date, a single meta-analysis dealing with objective assessment of knowledge was published, revealing no significant differences (SMD 0.15 (95% CI −0.09; 0.39), p = 0.22) across a variety of different tests [9]. This is consistent with our overall result. However, the confidence interval in the meta-analysis is also wide, including both little and significant values.

We plan to continue monitoring the effectiveness of our JC using tests suggested in the literature. We also would like to develop a validated test battery tailored to the assessment of knowledge in EBM and statistics.

Many publications report findings derived from self-reported questionnaires, observing increases in enjoyment related to participation in JCs, improvements in critical analysis skills, more well-thought application of results from articles, as well as increases in both reading time and the amount of sources studied [7, 8, 14, 33, 34]. We observed similar trends in our JC; however, we would like to emphasize that these are subjective, self-reported data, and the effectiveness of JCs should not be assessed solely on this basis. As Linzer notes, there is no association between self-reported outcomes and objective results [14].

The issue of test validity further complicates evaluation. To date, no clear standard exists for assessing knowledge in evidence-based medicine and statistics [2]. A number of methods have been proposed by different authors, but these tools are yet to be tested on diverse cohorts [18, 20, 23, 35, 36].

In our work, optional free-text comments submitted by participants proved to be very helpful, shedding light on both advantages and drawbacks of the JC organization; we always try to listen to these comments and use them to improve the JC.

In line with the model proposed by Linzer et al. and based on our results, we hypothesize that participation in a JC impacts clinical decision-making in practice [14]. However, this educational format cannot be assessed as a standard educational program; moreover, some of the relevant outcomes are inherently difficult or impossible to quantify and measure, which forces reliance on surrogate endpoints.

In Supplement 1, we provide the tests we used in our JC, so that researchers may use them in their practice. Supplement 2 presents a table describing each JC meeting conducted by the moment of data collection. This may be used for assessing our study as well as in developing new curricula or adapting existing ones.

This study has several limitations. First, the closed and online format of the JC may have reduced participant engagement and deterred potential applicants. Second, the heterogeneous sample, consisting mainly of residents and practicing physicians, limits the generalizability of our findings and complicates interpretation of how this educational format may affect learners at different stages of training. Nevertheless, we attempted to assess effectiveness the dynamics of standardizes test scores as surrogate outcomes, which yielded promising results, although requiring further corroboration. Third, the use of modified tests may have compromised their validity, hence limiting our ability to draw firm conclusions on the effectiveness of our journal club. Moreover, the variation in difficulty across tests does not allow to draw conclusions about the learning dynamics. Fourth, relatively low attendance may have biased assessments due to inadequate knowledge acquisition, effectively working as latent confounder. Fifth, the use of self-reported questionnaires is associated with such problems as nonresponse bias and extreme response bias. Sixth, the composition of the control group changed across recruitment waves, as it was not possible to track applicants who did not gain admission. This may lead to selection and allocation bias since some control participants may have pursued training in other JCs or acquired EBM and statistical knowledge independently. Seventh, a relatively small sample size, which may have reduced the statistical power of the study and widened the confidence intervals, i.e., increasing uncertainty. At the same time, increasing the number of participants might pose challenges for maintaining learning quality, individualized attention, and effective group discussions [37]. Eighth, residual confounding cannot be excluded, even after adjustment for identified potential confounders.

## Conclusion

We believe journal clubs are a valuable complement to formal academic medical education. They help keep students motivated, foster communication in the medical community, especially between young doctors and practicing physicians, facilitate the acquisition of new knowledge and skills, and promote their application in practice. However, assessing the effectiveness of journal clubs as educational interventions remains methodologically challenging, and existing evidence is inconclusive or controversial. This underscores the need for further research, and calls for developing methodologies for organizing and conducting JC meetings, as well as validated tools for the objective evaluation of gained knowledge.

## Supporting information

full text in Supplementary 1

## Data Availability

All data produced in the present study are available upon reasonable request to the authors

## Declarations

### Ethical approval and consent to participate

The authors confirm that all procedures contributing to this study complied with the ethical standards of the relevant national and institutional committees on human experimentation and were conducted in accordance with the Declaration of Helsinki (1975), as revised in 2008. Ethical approval for the study was obtained from the local Ethics Committee of the Leningrad Regional Clinical Hospital, St. Petersburg, Russia. The written informed consent was obtained from all participants prior to testing (administered via Google Forms), where the study was explicitly described as a part of the journal club framework. At the interview stage (Step 3 of the admission process), participants reconfirmed their consent

### Consent for publication

Not applicable

### Availability of data and materials

The datasets analyzed during the study are not publicly available due to the inclusion of personal information. However, they may be obtained from the corresponding author upon reasonable request

### Competing interests

All authors declare no conflict of interest

### Funding

Not applicable

### Authors’ contributions

Nikita Burlov – corresponding author; chief investigator; head of the journal club; study design; data acquisition, analysis and interpretation; agreement to be accountable for all aspects of the study.

Matvei Baranovskii – data acquisition and analysis; text drafting; agreement to be accountable for all aspects of the study.

Elizaveta Burlova – data interpretation; critical revision of the study; agreement to be accountable for all aspects of the study.

Matvei Slavenko – text editing and proofreading; additional data analysis and interpretation; final approval.

Gleb Khrykov – text drafting; study design; data interpretation; final approval; agreement to be accountable for all aspects of the study.

## Acknowledgments

Not applicable

## Notes

### Competing Interest Statement

The authors have declared no competing interest.

### Author Declarations

Ethical approval for the study was obtained from the local Ethics Committee of the Leningrad Regional Clinical Hospital, St. Petersburg, Russia in 09.2020

