## Supplementary material for "Prospective study on the organization and efficiency of online journal club": full text in Supplementary 1

### Supplementary material. Tests of evidence-based medicine and statistics.

#### Complex test

**USMLE sample test questions step1 2020 (1 point for each answer)**

№1. A study is designed to evaluate the feasibility of acupuncture in children with chronic headaches. Sixty children with chronic headaches are recruited for the study. In addition to their usual therapy, all children are treated with acupuncture three times a week for 2 months. Which of the following best describes this study design?

A. Case-control
**B. Case series**
C. Randomized clinical trial

№2. A placebo-controlled clinical trial is conducted to assess whether a new antihypertensive drug is more effective than standard therapy. A total of 5000 patients with essential hypertension are enrolled and randomly assigned to one of two groups: 2500 patients receive the new drug and 2500 patients receive placebo. If the alpha is set at 0.01 instead of 0.05, which of the following is the most likely result?

**A. Significant findings can be reported with greater confidence**
B. The study will have more power
C. There is a decreased likelihood of a Type II error
D. There is an increased likelihood of a Type I error

**Fresno Test of Evidence Based Medicine**

Clinical scenarios: «You have just seen Lydia who recently delivered a healthy baby. She plans to breastfeed, but also wants to start oral contraception. You generally prefer to prescribe combination oral contraceptives (estrogen + progesterone) but you have been told that these might more negatively affect her breastmilk production than progesterone only pills»

№3. Write a focused clinical question for each of these patient encounters that will help you organize a search of the clinical literature for an answer and choose the best article from among those you find.

**Answer: 1 point for a PICO question (else 0)**

№4. Where might clinicians go to find an answer to questions like these? Name as many possible types or categories of information sources as you can.

**Answer: 6 points for 4 different types of sources (databases of original research, journals, textbook, systematic reviews, professional medical guidelines, synthesized summaries, medical website, General internet search or people), 4 for 3, 2 for 2 (else 0)**

**«Design and Interpretation of Clinical Trials course» Coursera platform (1 point for each answer)**

№ 5. True or False: The purpose of a non-inferiority trial is to determine whether a new treatment is at least as effective as an already established treatment.

**True**

№6. True or False: In an adaptive design, adaptations are not specified in the protocol in order to allow investigators maximum flexibility for adjusting study parameters throughout the trial.

**False**

### Modified SPLIT test
**(1 point for each answer)**

**№1. To increase study power, one should increase:**

**A. Sample size**

B. P value

C. Statistical test power

D. Internal validity of research

E. Specificity

**№2. Why isn't arithmetic mean a representative measure of central tendency in a skewed distribution?**

A. Because a skewed distribution does not have a measure of central tendency.

B. Because it is difficult to present graphically.

C. Because skewed distribution cannot be mathematically defined.

**D. Because a single extreme result can significantly alter the mean.**

E. Because calculating arithmetic mean includes squaring, which in case of skewed distribution disproportionally increases extreme values.

**№3. Why does a randomised controlled trial (RCT) provide better quality of evidence than a cohort study?**

A. RCT has a control group, and a cohort study does not.

B. RCT has balanced group sizes, and a cohort study does not.

C. RCT is cheaper than a cohort study.

D. RCT is shorter in duration than a cohort study.

**E. RCT controls exposure to investigated factor, and a cohort study does not.**

**№4. The guideline for reporting cohort study results is called:**

A. COCHRANE.

B. PRISMA.

C. STARD.

D. CONSORT.

**E. STROBE.**

**№5. You want to search databases for information on breast cancer. Searching the key word «breast» retrieved 4200 articles, and key word «cancer» retrieved 34150 articles. Which of the following statements is correct?**

A. Using those two key words and the operator AND will retrieve 38150 articles.

**B. By using the operator OR to combine «breast» and «cancer» will retrieve at least 4200 articles.**

C. Search strategy «breast NOT cancer» will retrieve more than 4200 articles.

D. Search strategy «cancer NOT breast» will retrieve 34150 articles.

E. By using the operator OR you will get fewer than 4200 articles.

**№6. Prevalence of disease in a population is best determined by:**

A. Randomized controlled trial (RCT).

B. Cohort study.

**C. Cross-sectional study.**

D. Qualitative study.
E. Retrospective study.

**№7. Prevalence of headache in one year on a sample of Split-Dalmatia County citizens was measured, and these are the results:**

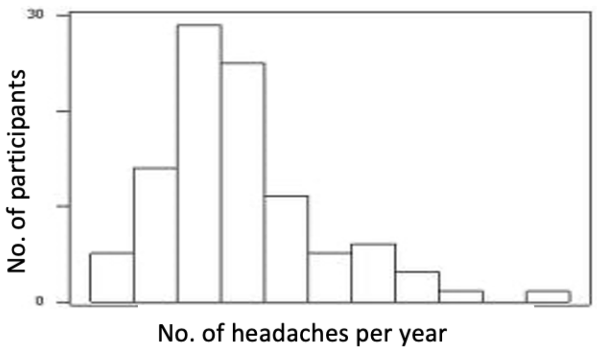

| | |
|---|---|
| Arithmetic mean | 16.8 |
| Median | 12 |
| Standard deviation | 2.79 |
| Interquartile range | 6 |
| Total range | 2-35 |

**Which measures should be used to describe the sample?**
A. Arithmetic mean and standard deviation (M=16.8, SD=2.79).
**B. Median and interquartile range (Md=12, IQR=6).**
C. Arithmetic mean and total range (M=16.8 and total range 2-35).
D. All of 5 listed measures, depending on statistical test being used.
E. Median and standard deviation (Md=12, SD=2.79).

**№8. The proportion of true negative results of a diagnostic test in the total number of healthy participants is called:**
A. Positive predictive value.
B. Precision.
C. Negative predictive value.
**D. Specificity.**
E. Sensitivity.

**№9. A new treatment for social anxiety was investigated and it provided normal everyday functioning in 20% of the patients, compared to 10% of the patients who were treated by standard treatment. How many patients should be included in the new program to assure that one of them achieves normal everyday functioning?**
A. 2.
B. 5.
C. 20.
**D. 10.**
E. 15.

**№10. A study of relation between consumption of black tea and weight loss in overweight participants gave the following results: group that consumed black tea for six months lost an**

**average of 328 g (95% CI 125 – 922 g), and the group that did not lost an average of 121 g (95% CI 50 – 211 g). What conclusions can you draw from this research?**

A. Black tea helped in reducing bodyweight for an average of 207 g.
B. Black tea helped in reducing bodyweight, but the result is not statistically significant.
C. Black tea helped in reducing bodyweight and the result is statistically significant.
**D. We cannot make conclusions about efficacy of black tea from available data.**
E. Black tea did not help in reducing bodyweight.

**№11. While investigating a possible connection between marijuana smoking and lung cancer, it was found that the number of participants who smoked marijuana was significantly higher in the group who had lung cancer than the healthy control group (OR=1.3, 95% CI 1.1-1.7). Correct interpretation of this result is:**

A. Subjects who smoke marijuana are 10-70% less likely to get lung cancer.
**B. Subjects who smoke marijuana are 30% more likely to get lung cancer.**
C. Subjects who smoke marijuana are 30% less likely to get lung cancer.
D. There is no association between marijuana smoking and lung cancer.
E. No conclusion can be made on the association between marijuana smoking and lung cancer.

**№12. A conclusion that participants exposed to a factor A had lower incidence of disease was made out of:**

A. Cross-sectional study.
**B. Cohort study.**
C. Diagnostic study.
D. Physiological study.
E. Case-control study.

**№13. The result of a case-control study is expressed as:**

**A. Odds ratio.**
B. Prevalence.
C. Absolute risk reduction.
D. Relative risk.
E. Positive predictive value.

**№14. Complete the summary with a conclusion.**

*Introduction:* To investigate if acute fear can actually «freeze the blood in your veins», researchers recruited 24 healthy volunteers (under 30 years).
*Methods:* Via random selection, 14 of the volunteers were sorted into a group that watched a horror movie before watching a documentary, and the remaining 10 volunteers watched the movies in reverse order. The movies were about 90 minutes long each and the volunteers watched them 7 days apart. Primary outcome was blood levels of coagulation factor VIII. Secondary outcome was participants' own experience of fear while watching the movies.
*Results:* Results show that, after watching the horror movie, mean difference in perceived fear was 5.4 units (95% CI 4.7-6.1) on a visual analogue scale (VAS), and blood levels of coagulation factor VII were significantly increased (mean difference 11.1 IU/dL (111 IU/L), 95% CI 1.2-21.0 IU/dL).

***Conclusion* is the following:**

A. There was a statistically significant difference in levels of coagulation factor VIII after watching the documentary.

**B. We cannot conclude with certainty, but the results indicate that horror movies increase blood levels of coagulation factor VII.**
C. There is not enough evidence to conclude that horror movies affect blood levels of coagulation factor VIII.
D. Fear can actually «freeze the blood in your veins».
E. There was a statistically significant increase of coagulation factor, but not of perceived fear.

**№15. You want to find the article that described the study mentioned above. What combination of key words would you choose to find the exact same article?**
A. Healthy volunteers, documentary/horror movie, factor VIII.
B. Healthy volunteers, factor VIII, visual analogue scale, horror movie.
C. Healthy volunteers, horror movie, visual analogue scale, factor VIII.
**D. Healthy volunteers, horror movie, documentary, factor VIII.**
E. Horror movie, documentary, visual analogue scale, factor VIII.

#### Assessing Competency in Evidence Based Medicine (ACE tool)

Read through the following information on patient scenario, clinical question, search strategy and article extract before answering the following set of questions.

| **Asking an answerable question** | **Yes** | **No** |
|---|---|---|
| 1. Are all PICO elements described in the patient scenario? | | |
| 2. Does the question constructed post-scenario provide a focused, foreground question? | | |
| **Searching the literature** | **Yes** | **No** |
| 3. Will the search strategy (to be used in Medline) retrieve relevant studies relating to the question? | | |
| 4. Does the search strategy utilise appropriate MeSH/keywords and Boolean operators correctly and effectively? | | |
| **Appraising the evidence** | **Yes** | **No** |
| 5. Was there sufficient information to determine the representativeness of the study participants? | | |
| 6. Was the method of participant allocation to intervention/exposure and comparison adequate? | | |
| 7. Was any form of adjustment required? | | |
| 8. Were all participants blinded to the treatment/exposure? | | |
| 9. Were all investigators blinded to the treatment/exposure? | | |
| 10. Were all outcome assessors blinded to the treatment/exposure? | | |
| 11. Were all patients analysed in the groups to which they were randomised? | | |
| **Applying the evidence** | **Yes** | **No** |
| 12. Does the patient in the scenario share similar characteristics/circumstances to participants in the study? | | |
| 13. Is the treatment/therapy feasible in the clinical setting of the scenario? | | |
| 14. Were all clinically important outcomes considered? | | |
| 15. Do the likely benefits of the treatment/therapy outweigh any potential harms and costs? | | |

**Patient scenario**

"Jane is a 42 year-old female Caucasian, who lives with her partner in metropolitan Melbourne, Australia. Jane is a lawyer, who quit smoking three years ago, after being a 'pack-a-day' smoker since her early 20s. Since her late 30s, Jane has received treatment for hypertension. Her medical history is otherwise unremarkable. At her most recent visit to her family doctor, Jane mentions that she has seen reports on the television about a new study investigating the preventive effects of aspirin. She has heard that aspirin may be beneficial in protecting against cardiovascular disease. Jane wonders whether she should be taking aspirin, given her history with hypertension, but wonders whether also being a diabetic might negate any benefit."

**Clinical Question**

"Is aspirin effective in reducing the risk of cardiovascular disease?"

**Search strategy**

| Item | Search | Results |
|---|---|---|
| 1 | Aspirin.mp | 52620 |
| 2 | exp Aspirin/ | 38658 |
| 3 | 1 OR 2 | 52620 |
| 4 | exp Cardiovascular Diseases/ | 1874575 |
| 5 | cardiovascular.mp. | 352938 |
| 6 | 4 OR 5 | 2003546 |
| 7 | Hypertension/ or hypertension.mp. | 355606 |
| 8 | diabetes.mp. or Diabetes Mellitus/ | 393986 |
| 9 | 3 AND 6 AND 7 AND 8 | 905 |
| 10 | Limit to randomised controlled trials | 75 |

**Article extract (hypothetical article)**

#### *A randomised controlled trial of aspirin for the prevention of cardiovascular disease*

##### *Background*

Aspirin is effective in the treatment of acute myocardial infarction and prevention of cardiovascular disease in men and women. Previous studies on the use of aspirin in primary prevention of cardiovascular disease have demonstrated a positive effect in men, yet the benefit in women remains uncertain. The aim of this study was to assess the effect of aspirin in the prevention of cardiovascular disease in women.

##### *Methods*

The study design was a randomised, double-blinded, placebo-controlled, trial of low-dose aspirin in the prevention of cardiovascular disease in women. The design of the study has previously been described in detail. In brief, between January 2002 and January 2012, letters of invitation were mailed to 500,000 women in the greater city of Melbourne, Victoria, Australia. A total of 63,250 volunteered to enrol in the study. Women were eligible if they were 40 years of age or older; had no history of coronary heart disease, cerebrovascular disease, no previous side-effects to taking aspirin and were not currently taking aspirin or any non-steroidal anti-inflammatory drug (NSAID) medication. A total of 31,150 women met the inclusion criteria of which 15,100 were randomised (through the generation of a computer generated scheme) to receive aspirin and 15,102 were randomised to receive the placebo. Written informed consent was obtained from all participants prior to commencement in the study. The trial was approved by the ethics board at the governing hospital and university institution. Participants in both groups were required to present every 6 months at the study site centre for assessment and to receive their medication. Medication was provided by the site pharmacy, which allocated identical appearing aspirin and placebo tablet in blister packs to the study's participants independent to the study's investigators. All participants were followed for myocardial infarction, stroke or death from cardiovascular causes. Medical records were obtained for all women in whom a cardiovascular event was recorded. These records were reviewed by an end-point committee, consisting of study investigators blinded to the treatment. The primary end point was cardiovascular events – a combination of myocardial infarction, stroke or death from cardiovascular causes. Only confirmed end-points of cardiovascular events were included in this study. Cox proportional hazard models were used to calculate hazard ratios and 95% confidence intervals for the comparison of event rates in the aspirin and placebo groups after adjustment for age.

##### *Results*

Both aspirin and placebo groups were similar with respect to baseline characteristics (Table 1). The average duration of follow-up from randomisation to the end of the trial was 4.2 years (range, 2.3 to 5.0 years). Throughout the duration of the trial, drop-outs occurred. Data presented is based on participants that completed the trial during the study period. A total of 422 women in the aspirin group and 478 women in the placebo group had a cardiovascular event (Hazard Ratio, 0.83; 95% confidence interval, 0.77 to 1.01). There was no evidence that any of the cardiovascular risk factors considered, except smoking status and hyperlipidemia, modified the effect of aspirin on the primary end-point.

##### *Discussion*

In this large study, involving 63,250 women, a 100 mg daily dose of prophylactic aspirin is associated with a reduced risk of major cardiovascular events. No significant evidence was found that age, hypertension, diabetes or BMI modified the effect of aspirin. Middle aged women who adhere to a daily low dose of aspirin can significantly reduce the risk of cardiovascular disease. The rate of benefit is large, with a cardiovascular event prevented for every 269 women treated with aspirin.

| Table 1. Baseline characteristics | | | |
|---|---|---|---|
| | Aspirin (N=15,100) | Placebo (N=15,102) | Total (N=30,202) |
| Age (years) | | | |
| (mean±SD) | 55.3±8.0 | 54.9±8.0 | 55.1±8.0 |
| 40-50 (%) | 50.2 | 50.1 | 50.1 |
| 51-60 (%) | 42.9 | 43.0 | 43.0 |
| >61 (%) | 6.9 | 6.9 | 6.9 |
| Smoking status | | | |
| Current (%) + | 15.0 | 14.7 | 14.9 |
| Past/never (%) | 85.0 | 85.3 | 85.1 |
| Body mass index ($kgm^{-2}$) | | | |
| (mean±SD) | 25.1±4.3 | 25.3±4.3 | 25.2±4.3 |
| ≤25.0 (%) | 48.8 | 48.8 | 48.8 |
| 25.1-29.9 (%) | 32.1 | 32.2 | 32.2 |
| ≥30.0 (%) | 19.1 | 19.0 | 19.0 |
| Hypertension | | | |
| Yes (%) | 25.0 | 24.9 | 25.0 |
| No (%) | 75.0 | 75.1 | 75.0 |
| Diabetes | | | |
| Yes (%) | 2.3% | 2.2% | 2.2% |
| No (%) | 97.7% | 97.8% | 97.8% |
| Hyperlipidemia | | | |
| Yes (%) | 27.3 | 27.2 | 27.2 |
| No (%) | 72.7 | 72.8 | 72.8 |

Mean differences tested using independent t-test; proportional differences tested using the chi square test. +significantly different at $p<0.05$.

| Table 2. Hazard ratios of cardiovascular events, related to baseline characteristics | | | | |
|---|---|---|---|---|
| | Total number | Aspirin | Placebo | HR (95% CI) |
| Age (years) | | | | |
| 40-50 | 15131 | 122 | 142 | 0.86 (0.67-1.09) |
| 51-60 | 12987 | 148 | 166 | 0.89 (0.71-1.13) |
| >61 | 2084 | 152 | 170 | 0.90 (0.74-1.11) |
| Smoking status | | | | |
| Current | 4500 | 159 | 140 | 1.12 (1.00-1.40) |
| Past/never | 25702 | 263 | 338 | 0.78 (0.66-0.92) |
| Body mass index ($kgm^{-2}$) | | | | |
| ≤25.0 | 14738 | 181 | 208 | 0.87 (0.71-1.06) |
| 25.1-29.9 | 9725 | 150 | 169 | 0.97 (0.71-1.11) |
| ≥30.0 | 5739 | 91 | 101 | 0.90 (0.68-1.20) |
| Hypertension | | | | |
| Yes | 5051 | 221 | 250 | 0.89 (0.75-1.06) |
| No | 25151 | 201 | 228 | 0.87 (0.73-1.06) |
| Diabetes | | | | |
| Yes | 664 | 58 | 62 | 0.94 (0.68-1.31) |
| No | 29538 | 364 | 416 | 0.87 (0.76-1.01) |
| Hyperlipidemia | | | | |
| Yes | 8214 | 196 | 168 | 1.15 (1.04-1.48) |
| No | 21988 | 226 | 310 | 0.73 (0.62-0.87) |

#### Assessing Competency in Evidence Based Medicine (ACE tool)

**Answers**

| **Asking an answerable question** | **Yes** | **No** |
|---|---|---|
| 1. Are all PICO elements described in the patient scenario? | ✓ | |
| 2. Does the question constructed post-scenario provide a focused, foreground question? | | ✓ |
| **Searching the literature** | **Yes** | **No** |
| 3. Will the search strategy (to be used in Medline) retrieve relevant studies relating to the question? | ✓ | |
| 4. Does the search strategy utilise appropriate MeSH/keywords and Boolean operators correctly and effectively? | ✓ | |
| **Appraising the evidence** | **Yes** | **No** |
| 5. Was there sufficient information to determine the representativeness of the study participants? | ✓ | |
| 6. Was the method of participant allocation to intervention/exposure and comparison adequate? | | ✓ |
| 7. Was any form of adjustment required? | | ✓ |
| 8. Were all participants blinded to the treatment/exposure? | ✓ | |
| 9. Were all investigators blinded to the treatment/exposure? | ✓ | |
| 10. Were all outcome assessors blinded to the treatment/exposure? | ✓ | |
| 11. Were all patients analysed in the groups to which they were randomised? | | ✓ |
| **Applying the evidence** | **Yes** | **No** |
| 12. Does the patient in the scenario share similar characteristics/circumstances to participants in the study? | ✓ | |
| 13. Is the treatment/therapy feasible in the clinical setting of the scenario? | ✓ | |
| 14. Were all clinically important outcomes were considered? | | ✓ |
| 15. Do the likely benefits of the treatment/therapy outweigh any potential harms and costs? | | ✓ |

**Reasoning for answers**

1. All PICO elements are provided in the patient scenario.
2. PICO question is not focused (“Is aspirin effective in reducing the risk of cardiovascular disease?”), no mention of patient aspects only intervention and outcome.
3. Search strategy will retrieve relevant studies.
4. Appropriate use of MeSH/keywords and Boolean operators.
5. Information regarding source, eligible and participant population was made available.
6. Randomisation is mentioned, but no detail on how this randomisation process was achieved or if allocation concealment was achieved.
7. No adjustment necessary as baseline characteristics are similar.
8. Participants were blinded to treatment (use of pharmacy to dispense identical looking pills).
9. Investigators blinded through use of third party (pharmacy) to dispense intervention and placebo tablets.
10. Outcomes assessed by end-point committee, consisting of study investigators blinded to the treatment.
11. Drop-outs occurred, no ITT analysis performed, analysis only on participants completing the trial (per protocol analysis).
12. Age, smoking status, hypertension and diabetes all relevant to the patient, and reported in the study.
13. Treatment (aspirin) is feasible in the clinical setting (i.e. general practice, metropolitan Melbourne).
14. Clinically important outcomes such as side effects of aspirin were not reported.
15. Hazard ratio does not report a statistically significant decrease in the risk of cardiovascular events. No evidence on potential harms, or costs.

### Berlin test

**(1 point for each answer)**

QUESTION 1

A man arrives in the emergency room with pain in his right lower abdomen area that has been present for about 24 hours. There are no clear signs of appendicitis on clinical examination. However, you know that in the patient's age group, approximately every tenth patient with these symptoms has appendicitis without typical signs.

You initiate an ultrasound examination because you know from the last internal quality control that the sonographer on duty has achieved good results in appendicitis diagnostics (probability ratio [= likelihood ratio] for positive findings 1.8, for negative results 0.2).

In this case, the sonographer determines that the patient has appendicitis. When you telephone the surgeon, he asks you what the likelihood is that the patient actually has appendicitis.

You answer:

A. About 2%
B. About 7%
C. About 15%
D. About 30%
E. A statement is not possible before the laboratory findings arrive (An unlabelled Fagan nomogram is included)

**Answer 1: __C__**

QUESTION 2

The trainee who is present is impressed with your answer and asks you how to calculate a probability ratio. You tell him that the ultrasound diagnoses from a specific period of time are compared to the actual correct diagnoses (from histology or follow-up). Since you do not remember the numbers, demonstrate this using a numerical example:

| | | Actual diagnosis of appendicitis | |
|---|---|---|---|
| | | Yes | No |
| Sonographic diagnosis | Yes | 90 | 10 |
| appendicitis | No | 20 | 110 |

In this example, the probability ratio for a positive finding is:

**A** $$0{,}09 = \frac{\frac{10}{(10+110)}}{1 - \frac{20}{(110+90)}}$$

**B** $$0{,}10 = \frac{\frac{10}{(110+10)}}{\frac{90}{(90+20)}}$$

**C** $$1{,}54 = \frac{\frac{20}{(20+110)}}{1 - \frac{90}{(90+10)}}$$

**D** $$9{,}82 = \frac{\frac{90}{(90+20)}}{\frac{10}{(10+110)}}$$

E. The probability ratio cannot be calculated from this information

**Answer 2: __D____**

QUESTION 3

The trainee is now very enthusiastic, claiming more about the patients in the numerical example (question 2):

1) Without any further information, it can be stated with a 90 per cent probability that the sample patients with pathological ultrasound findings have appendicitis.
2) Without any further information, it can be stated that for the sample patients with normal ultrasound findings the probability of a false finding is 18% → [20÷(20+90)].
3) The positive predictive value is calculated directly from the quotient of the probability ratios → [90%=0.09÷0.01].

You tell him:

A. All statements (1, 2, 3) are incorrect.
B. The first statement (1) is correct; the other statements (2, 3) are incorrect.
C. The second and third statements (2, 3) are correct; the first statement (1) is incorrect.
D. The first and third statements (1, 3) are correct; the second statement (2) is incorrect.
E. All statements (1, 2, 3) are correct.

**Answer 3: __A____**

QUESTION 4

A pharmaceutical representative visit you in your practice and introduces you to a new drug that in a large double-blind randomised controlled study on healthy employees of the pharmaceutical company - has achieved a fifty per cent reduction in the risk of dying of a heart attack:

- 4,000 people were treated, of whom 4 (0.1%) died from heart attack.
- Of the 4,000 untreated controls, 8 (0.2%) died.

The pharmaceutical representative, therefore, recommends treating all patients with the new drug. You want to save the life of at least one patient in your practice, but find that you have to treat the following number of patients to achieve this:

A. 1,000 patients [= 1÷(0.2%-0.1%)]
B. 2,000 patients [= 8,000÷4]
C. 4,000 patients [= 4 •(1÷0.1%)]
D. 8,000 patients [= 4,000•2]
E. cannot calculate the number of patients that you need to treat

**Answer 4: __A____**

QUESTION 5

You enjoy this opportunity to help humankind and call a colleague to share the good news. Your colleague has already read the publication of the study data (question 4) and points out the following number of cases of pulmonary oedema of unclear origin:

- 7 in the treatment group
- 2 in the control group

You check and find that one additional pulmonary oedema can be expected when the following number of patients is treated:

A. 1,000 treated patients [= 2,000*0.2%]

B. 571 treated patients [= 4,000÷7]
C. 800 treated patients [= 1÷(5÷4,000)]
D. 2,000 treated patients [= 2*(1÷0.1%)]
E. The number of patients to be treated cannot be calculated from this data.

**Answer 5: __C__**

QUESTION 6

A patient with chronic headache recently read in her TV magazine that exposure to formaldehyde could cause chronic headache. She now wants the health insurance to fund her move. Therefore, she asks whether connection between chronic headache and formaldehyde in indoor air has been scientifically proven. You do a literature search and find various studies on this topic.

Which study design do you think would be the most appropriate for studying this question?

A. Prevalence study
B. Ecological study
C. Case-control study
D. Prospective randomised controlled study
E. Case report

**Answer 6: __B__**

QUESTION 7

In detail, the following procedures have been chosen for the studies you have found. Which do you think would be the best way to check whether there is a connection between headache and formaldehyde in the room?

A. Measure the formaldehyde content in the living quarters of 100 patients from a specialised ambulance who have a headache and 100 patients from general dental practices without a headache, but who correspond to the headache patients in age, gender and income.
    - Compare the mean formaldehyde content in both groups.
B. Interview 500 patients at an environmental medicine ambulance, and ask whether they have a headache and whether their homes are exposed to formaldehyde.
    - Compare the frequencies of the two answers (formaldehyde-stressed living rooms) from patients with a headache vs patients without a headache.
C. Interview tenants from a housing company using a headache questionnaire. Simultaneously perform a skin test for formaldehyde allergy.
    - Compare the incidence of headache in tenants with formaldehyde allergy vs remaining tenants.
D. Perform a two-time measurement of the formaldehyde concentration in the blood of headache patients from a pain clinic at a one-year intervals.
    - Compare the values at the beginning with those at the end of the observation.
E. Survey, using a headache questionnaire, persons who have newly moved into presumably formaldehyde-contaminated living quarters of an urban housing company and long-term tenants of such apartments.
    - Compare the frequency of headache between new and long-term tenants.

**Answer 7: B**

QUESTION 8

A large study has investigated whether a new slimming pill reduces mortality from cardiovascular mortality. The study involved 1,000 high-overweight patients. At random, 500 patients were placed in the treatment group and 500 in the control group. The treatment group received the slimming pills for over a year, while the control group received a similar-looking lactose pill (placebo). The patients in the treatment group were examined monthly for the occurrence of side effects while the patients in the control group were examined only twice a year.
In the treatment group, the number of patients who died of cardiovascular mortality was 10 less than in the control group ($p < 0.01$). Consider whether the quality of the study will convince you. The following statement is most likely to apply:

A. The study was not conducted prospectively.
B. The study was not randomised.
C. The study was not performed double-blind.
D. The study did not investigate the issue endpoint (which was interesting for the study) but rather a surrogate endpoint.
E. In this prospective randomised-controlled double-blind study, the endpoint of interest for the research question was investigated.

**Answer 8: C**

QUESTION 9

1) From the available information (question 8), it is possible to calculate how many patients, as in the study, have to be treated with the slimming pills in order to prevent an additional cardiovascular death.
2) In order to calculate the number of patients that need to be treated to prevent an additional death, one must know the relative reduction in the risk of death of the treatment group over the control group.
3) The available data (question 8) indicate the relative reduction in the risk of death of the treatment group compared to the control group (relative risk reduction).

A. All statements (1, 2, 3) are incorrect.
B. The first statement (1) is correct; the other statements (2, 3) are incorrect.
C. The second and third statements (2, 3) are correct; the first statement (1) is incorrect.
D. The first and third statements (1, 3) are correct; the second statement (2) is incorrect.
E. All statements (1, 2, 3) are correct.

**Answer 9: B**

QUESTION 10

You are a internist. Your 63-year-old female patient has a 70% stenosis of the carotid artery as an incidental finding. You are wondering whether this is an indication for referral for endarterectomy. You find a study that shows no benefit from surgery for conservative treatment in asymptomatic patients with 70% stenosis (comparable to your patient) at 5 years follow-up. In

a subgroup analysis (13 subgroups in total), after taking into account the risk factor status at baseline, only those women who survived the first year without insult or TIA showed over the following 4 years a statistically significant benefit ($p < 0.03$) from endarterectomy.
Which statement is correct?

A. The significant result demonstrates a benefit to women and is sufficient alone to justify the indication for surgery.
B. In the subgroup, analysis has been corrected for other risk factors, which usually leads to misleading conclusions.
C. Subgroup analyses maximise the yield of reliable results from randomised controlled trials.
D. As the number of subsequently formed subgroups increases, there is an increased danger that a subset will erroneously find a benefit that does not exist.
E. The subgroup analysis, in this case, shows that asymptomatic stenosis of the carotid artery is more dangerous for women than for men.

**Answer 10: __D_____**

QUESTION 11

In an education course on the effects of lipid reducers after myocardial infarction, the following randomised controlled trials are presented. Various medications were used in them. All studies included a placebo group with middle-aged (55-year-old) patients with adjusted hypertension (RR: 155/98 mmHg) and obesity. The medications were tested against a placebo over a period of 5 years.

1) In a Bolivian study, therapy II reduced the risk of heart attack by 25%.
2) In an Argentinian study, therapy I lowered the risk of deathly infarction by 30%.
3) In a Chilean study, 3% of the patients in the treatment III group and 4% in the control group died from heart attack.

Which statement is correct?

A. Therapy 1 is preferred because this therapy reduces the risk of heart attack the most.
B. Therapy 3 is preferred because the majority of her patients benefit from the treatment.
C. For therapies 2 and 3, the relative reduction in the risk of heart attack is the same.
D. For therapies 1 and 3, the relative reduction in the risk of a deathly infarction is the same.
E. None of the therapies indicates the risk for untreated patients (control event rate).

**Answer 11: __A_____**

QUESTION 12

In a gastro-enterological policlinic, the prevalence of colon cancer is 30%. One thousand consecutive patients will be included in a study for a new, non-invasive, low-stress diagnostic test for the detection of colon cancer. The test detects 630 patients as being truly negative, properly tumor free. The number of false-negative and false-positive patients is identical.

| | Gold standard positive | Gold standard negative |
|---|---|---|
| Test positive | | |
| Test negative | | |

(4-field table for your own calculation of the result)

Which 4-field table corresponds to this information?
Four-field-Table A

| | Gold standard positive | Gold standard negative | |
|---|---|---|---|
| Test positive | 230 | 70 | 300 |
| Test negative | 70 | 630 | 700 |
| | 300 | 700 | 1000 patients |

Four-field-Table B

| | Gold standard positive | Gold standard negative | |
|---|---|---|---|
| Test positive | 300 | 0 | 300 |
| Test negative | 0 | 700 | 700 |
| | 300 | 700 | 1000 patients |

Four-field-Table C

| | Gold standard positive | Gold standard negative | |
|---|---|---|---|
| Test positive | 270 | 100 | 370 |
| Test negative | 30 | 600 | 630 |
| | 300 | 700 | 1000 patients |

Four-field-Table D

| | Gold standard positive | Gold standard negative | |
|---|---|---|---|
| Test positive | 670 | 30 | 700 |
| Test negative | 30 | 270 | 300 |
| | 700 | 300 | 1000 patients |

Four-field-Table E

| | Gold standard positive | Gold standard negative | |
|---|---|---|---|
| Test positive | 300 | 70 | 370 |
| Test negative | 70 | 560 | 630 |
| | 370 | 630 | 1000 patients |

**Answer 12: __A_____**

QUESTION 13

Some time ago, you referred a 40-year-old female patient to a gynaecologist because of a palpable nodule in the breast. The nodule was classified as benign after the puncture. The patient brings her findings and wants to know whether the cyst means she's at particular risk of developing breast cancer. You find several studies. Which study is most suitable for assessing the prognostic significance of benign cysts in patients from a normal population?

A. A study from the Gynaecologist Clinic of a Paris university hospital: From 1996−1998. All patients with carcinoma who were found to have palpable cysts were interviewed. For comparison, patients without carcinoma were examined for palpable cysts. Patients with carcinoma had 20% fewer cysts than patients without carcinoma.

B. A study from a university hospital in Boston: mammograms (X-rays) from 1,500 patients spanning the last 10 years and showing carcinoma retrospectively examined for cysts. Thirty per cent of the patients also had larger cysts.
C. A study from the gynaecological ambulance of the only referral centre in Eastern Scotland for patients with problems in the breast: All patients with palpable cysts were examined 10 years later or followed through the breast cancer registry and death records. Compared with the normal population, the frequency for carcinomas was twice as high.
D. A study from the pathology tissues at a special clinic in the Ruhr area: All tissue histology from the last 10 years was recorded. Eleven per cent of patients with one benign cyst were also diagnosed histologically with carcinoma within the detection period.
E. A multicentre study in several district hospitals: Experienced surgeons were asked how often carcinoma patients were diagnosed with benign cysts. The mean of the information was 37%.

**Answer 13: C**

QUESTION 14

In German-speaking countries, a free-floating thrombus is particularly feared in deep leg vein thrombosis because of the presumed higher embolic risk. You would like to know whether patients with a free-floating thrombus are at a higher risk for pulmonary embolism than patients with a wall-mounted thrombus.

1) This is a question about a forecast.
2) This is a question about side effects.
3) This question is best examined in a case- control study.
4) This question is best studied in a cohort study.
5) This question is best studied in a randomised controlled trial.

Which statement is correct?

A. 1 and 3 are correct
B. 1 and 4 are correct
C. 1 and 5 are correct
D. 2 and 4 are correct
E. 2 and 5 are correct

**Answer 14: B**

QUESTION 15:

A meta-analysis is a type of study that has gained increasing importance lately. Which statement is correct?

A. Through the technique of meta-analysis, large studies with many patients (mega-trials) have become less significant.
B. Systematic studies have shown that studies conducted in the respective national language are worse than the studies in English magazines. For this reason, it is sufficient for a meta-analysis to consider the view of English-language literature.
C. Studies with larger numbers of patients often find a greater therapeutic effect than studies with a smaller number of patients.

D. In a meta-analysis of randomised trials with a high number of patients, the effectiveness of measures can be estimated more accurately than in smaller individual studies.
E. In a meta-analysis, there may be differences in the question or the patient populations of the individual studies may be balanced by using statistics.

**Answer 15: __D_____**

### RESET test

**(The remaining items are given one point if the correct option was selected and no points otherwise. Questions represented four EBM constructs: ask (Q1), appraise (Q2–Q7), apply (Q9, Q13, Q15–Q18), and interpreting results (Q8, Q10–Q12, Q14). The questions were weighted by multiplying by the following scalar: ask (3) appraise (2) apply (2) and interpreting results (2.4) and then summed to obtain the overall score)**

1. You are seeing an 80 year-old male who has a history of heart failure, stroke and atrial fibrillation. He is currently on warfarin and his INR is therapeutic 70% of the time. You recently heard about a new medication, dabigatran, which does not require monitoring of INR.

Write a structured clinical question to address one aspect of this clinical encounter.

**1 point for each components PICO question**

2. You are considering using an antibiotic, Killamicin, for a patient of yours with MRSA. Which of the following would offer you the best evidence of the antibiotic's efficacy?

A. a meta-analysis of 14 trials (2,012 patients) published in *JAMA* in 2002
B. a retrospective cohort study of 25,000 patients published in *Archives of Internal Medicine* in 2004
C. a narrative review published in *NEJM* in 2008
**D. a randomized trial of 2,000 patients published in *Clinical Infectious Diseases* in 2006**

3. In a randomized controlled trial of statins for primary and secondary prevention, 70% of those randomized to statins took the medication. Of those randomized to placebo, 20% received prescriptions for a statin (either the study drug or another statin) from their doctor. What is the appropriate way to analyze this study to minimize bias?

A. Compare the outcomes of patients who took statins to those who did not, adjusting for any baseline differences between the two groups.
**B. Compare the outcomes of those randomized to statins to those randomized to placebo, regardless of what they took.**
C. Compare the outcomes of patients randomized to statins who took statins to those randomized to placebo who took the placebo.
D. Compare the outcomes of patients randomized to statins who took statins to those randomized to placebo, regardless of what they took.

4. The Nurses' Health Study was a prospective cohort study enrolling 121,700 female nurses, 20 to 55 years of age. Every 2 years, participants completed a mailed questionnaire about their use of postmenopausal hormones and their medical history, including cardiovascular disease and associated risk factors. Follow-up ended when cardiovascular disease was first diagnosed, the participant died, or the last questionnaire was returned.
The primary outcome was cardiovascular disease. Proportional-hazards models were used to calculate relative risks, with adjustments for age, body-mass index, cigarette smoking, hypertension, diabetes, elevated cholesterol levels, and family history.

All of the patients were Caucasian. Non-users of hormone therapy were older and were twice as likely to smoke as hormone users. Hormone users had an adjusted risk of MI that was 60% that of non-users.

Which of the following is most true about the internal validity of this study?

**A. The adjusted results are not confounded by smoking status.**
B. The adjusted results are confounded by age.
C. The adjusted results demonstrate selection bias because all of the participants were Caucasian.
D. The adjusted results are invalid due to recall bias.

5. In a case-control study to assess the effect of thiazide diuretics on sudden cardiac death, cases of hypertensive patients with sudden cardiac death are identified from a death registry. What is the appropriate control population?

A. Age-matched patients who died of other causes.
B. Age-matched patients on loop diuretics.
**C. Age-matched patients with hypertension.**
D. Age-matched patients with coronary artery disease.

*Siscovick et al. Diuretic Therapy for Hypertension and the Risk of Primary Cardiac Arrest. N Engl J Med 1994; 330:1852-1857*

6. Which of the following is true of randomized controlled trials?

A. They are not subject to crossovers.
B. They may be subject to selection bias.
**C. Their results may be used to calculate an absolute risk reduction.**
D. Their validity is not affected by loss to follow-up.

7. Which of the following is NOT true of a cohort study?

A. It can be retrospective or prospective.
**B. It can establish cause and effect.**
C. It may be subject to selection bias.
D. It often includes “real world” patients.

8. A colleague has developed a blood test which she feels can screen for cervical cancer in females. She performed her test on a number of patients with and without cervical cancer and produced the following results:

| | | Cervical Cancer (as determined by the Gold Standard) | |
|---|---|---|---|
| | | Cancer | No Cancer |
| Cervical Cancer Blood Test Result | Positive | 20 | 20 |
| | Negative | 10 | 80 |

Based on the table, what is the ***specificity*** of the cervical cancer blood test?

A. 20%
B. 50%
C. 67%
**D. 80%**

9. According to PIOPED II, CT pulmonary angiography has a sensitivity of 83% and a specificity of 96%. If a patient has a positive CT pulmonary angiogram, what is the post-test probability that he has a pulmonary embolism?

A. 83%
B. 96%
C. 99%
**D. cannot be calculated from the information given**

*Stein et al. Multidetector computed tomography for acute pulmonary embolism. N Engl J Med. 2006;354(22):2317*

(Questions 10-12) You have recently finished reading a study describing how a new antiviral as compared to Vitamin C can prevent the common cold. The results of the study are depicted below.

| | New Antiviral (n = 200) | Vitamin C (n = 200) |
|---|---|---|
| Primary Outcome: PCP visit for cold symptoms | 60 (30%) | 80 (40%) |

10. What is the absolute risk reduction of the primary outcome?

**A. 10%**
B. 20%
C. 25%
D. 75%

11. What is the relative risk reduction of the primary outcome?

A. 10%
B. 20%
**C. 25%**
D. 75%

12. What is the number needed to treat?

A. 4
**B. 10**
C. 20
D. 75

13. You are seeing a 50 year-old gentleman with a history of CAD and well-controlled depression for smoking cessation. You and your patient are discussing treatment modalities to help him quit. You are considering giving your patient varenicline and have reviewed the recent pertinent literature. A randomized controlled trial in 2008 compared varenicline to bupropion. In addition to drug therapy, all participants were given a smoking cessation self-help booklet and were provided with individual counseling during weekly follow-up visits for 12 consecutive weeks. Because varenicline has been associated with neuropsychiatric side effects, patients with major depression were excluded from the study. Based on this trial, you understand that varenicline was more effective than bupropion. You are able to provide your patient with varenicline but are unable to provide him with weekly counseling visits or a self-help booklet.

*Gonzales D, et al. Varenicline, an alpha4beta2 nicotinic acetylcholine receptor partial agonist, vs sustained-release bupropion and placebo for smoking cessation: a randomized controlled trial. JAMA. 2006;296(1):47.*

Which of the following statements is most likely to be true regarding the use of varenicline in this patient?

A. Varenicline would be less effective in your patient than in the trial participants because he would know that he is receiving varenicline (no placebo effect).
**B. Varenicline would cause your patient to experience greater neuropsychiatric risk because your patient has a history of depression.**
C. Varenicline would not be effective for your patient because he would have been excluded from the study.
D. Varenicline would be as effective in your patient because behavioral counseling is not an important aspect of smoking cessation care.

14. You are reviewing a recent article about a superstatin that has been shown to decrease mortality in high-risk cardiovascular patients. The 1-year all-cause mortality rate was 5% in patients who were given the test drug and 10% in patients receiving placebo. The p-value was 0.06 and the 95% confidence interval for the absolute risk reduction was -1% to 11%.

Which of the following is true?

A. The superstatin is not effective because the p-value is greater than 0.05.
B. There is a 94% chance that the superstatin reduces all-cause mortality by 5%.
**C. The superstatin might be effective but a larger study is necessary to prove whether it is or is not.**
D. The superstatin is not effective because the confidence interval crosses 1.

15. The CURE trial sought to determine whether aspirin + placebo or aspirin + clopidogrel were more effective in preventing death in patients who presented within 24 hours of a NSTEMI. Of the following patients (see forest plot below), which group of patients would **<u>not</u>** benefit from clopidogrel in addition to aspirin?

*Yusuf S, et al Effects of clopidogrel in addition to aspirin in patients with acute coronary syndromes without ST-segment elevation. N Engl J Med. 2001;345(7):494.*

A. Patients over age 65 years
B. Patients with no history of revascularization
C. Female patients
D. Patients without diabetes
**E. All of these groups will likely benefit**

| Characteristic | No. of Patients | Percentage of Patients with Event: Placebo | Percentage of Patients with Event: Clopidogrel |
|---|---|---|---|
| Overall | 12562 | 11.4 | 9.3 |
| Associated MI | 3283 | 13.7 | 11.3 |
| No associated MI | 9279 | 10.6 | 8.6 |
| Male sex | 7726 | 11.9 | 9.1 |
| Female sex | 4836 | 10.7 | 9.5 |
| ≤65 yr old | 6354 | 7.6 | 5.4 |
| >65 yr old | 6208 | 15.3 | 13.3 |
| ST-segment deviation | 6275 | 14.3 | 11.5 |
| No ST-segment deviation | 6287 | 8.6 | 7.0 |
| Enzymes elevated at entry | 3176 | 13.0 | 10.7 |
| Enzymes not elevated at entry | 9386 | 10.9 | 8.8 |
| Diabetes | 2840 | 16.7 | 14.2 |
| No diabetes | 9722 | 9.9 | 7.9 |
| Low risk | 4187 | 6.7 | 5.1 |
| Intermediate risk | 4185 | 9.4 | 6.5 |
| High risk | 4184 | 18.0 | 16.3 |
| History of revascularization | 2246 | 14.4 | 8.4 |
| No history of revascularization | 10316 | 10.7 | 9.5 |
| Revascularization after randomization | 4577 | 13.9 | 11.5 |
| No revascularization after randomization | 7985 | 10.0 | 8.1 |

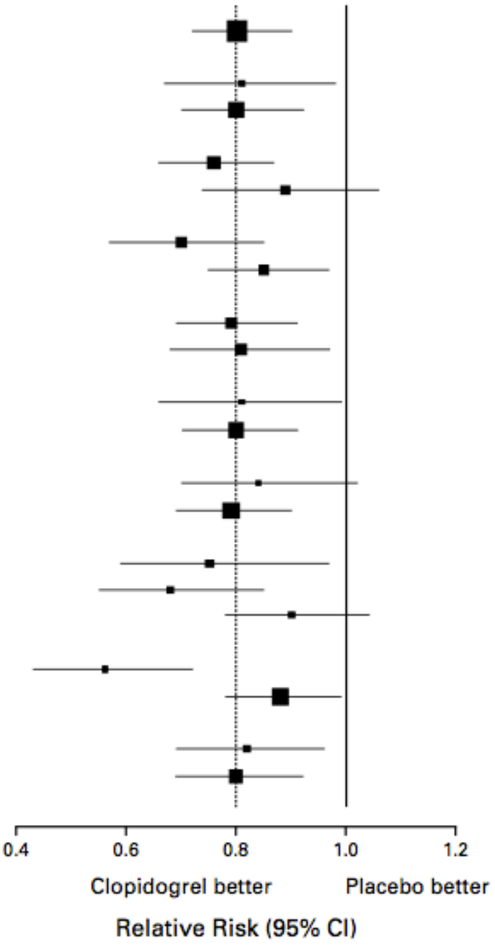

16. You are admitting a 75 year-old female for knee pain. She has a history of diabetes and a knee prosthesis. Based on your history and clinical exam, you suspect that the pre-test probability of this patient having septic arthritis is 90%. An ESR was ordered and returns at 25 mm/hr. What is the post-test probability that your patient has septic arthritis?

**Table 2.** Likelihood Ratios for Risk Factors, Signs, and Serum Laboratory Values

| | Source | Sensitivity, % | Specificity, % | Relative Risk | Likelihood Ratio (95% CI): Positive | Likelihood Ratio (95% CI): Negative |
|---|---|---|---|---|---|---|
| Risk factors | | | | | | |
| Age >80 y | Kaandorp et al,[42] 1995 | 19 | 95 | 4.1 | 3.5 (1.8-7.0) | 0.86 (0.73-1.00) |
| Diabetes mellitus | Kaandorp et al,[42] 1995 | 12 | 96 | 2.8 | 2.7 (1.0-6.9) | 0.93 (0.83-1.00) |
| Rheumatoid arthritis | Kaandorp et al,[42] 1995 | 68 | 73 | 5.4 | 2.5 (2.0-3.1) | 0.45 (0.32-0.72) |
| Recent joint surgery | Kaandorp et al,[42] 1995 | 24 | 96 | 8.4 | 6.9 (3.8-12.0) | 0.78 (0.64-0.94) |
| Hip or knee prosthesis | Kaandorp et al,[42] 1995 | 35 | 89 | 4.1 | 3.1 (2.0-4.9) | 0.73 (0.57-0.93) |
| Skin infection | Kaandorp et al,[42] 1995 | 32 | 88 | 3.6 | 2.8 (1.7-4.5) | 0.76 (0.60-0.96) |
| Hip or knee prosthesis and skin infection | Kaandorp et al,[42] 1995 | 24 | 98 | 18 | 15.0 (8.1-28.0) | 0.77 (0.64-0.93) |
| HIV-1 infection | Saraux et al,[43] 1997 | 79 | 50 | 3.2 | 1.7 (1.0-2.8) | 0.47 (0.25-0.90) |
| Physical examination | | | | | | |
| Fever | Kortekangas et al,[47] 1992 | 46 | 31 | NA | 0.67 (0.43-1.00) | 1.7 (1.0-3.0) |
| Serum laboratory values* | | | | | | |
| Abnormal peripheral WBC count | Jeng et al,[48] 1997 | 90 | 36 | NA | 1.4 (1.1-1.8) | 0.28 (0.07-1.10) |
| Erythrocyte sedimentation rate | Jeng et al,[48] 1997 | 95 | 29 | NA | 1.3 (1.1-1.8) | 0.17 (0.20-1.30) |
| C-reactive protein | Söderquist et al,[44] 1998 | 77 | 53 | NA | 1.6 (1.1-2.5) | 0.44 (0.24-0.82) |

Abbreviations: CI, confidence interval; HIV-1, human immunodeficiency virus type 1; NA, not applicable; WBC, white blood cell.
*Defined as abnormal peripheral WBC count of more than 10 000/μL, elevated erythrocyte sedimentation rate of more than 30 mm/h, and elevated C-reactive protein of more than 100 mg/L.

*Margaretten, ME, et al. Does this adult patient have septic arthritis? JAMA. 2007;297(13):1478-88.*

A. 15%
**B. 60%**
C. 86%
D. 92%

17. Below is a Kaplan-Meier curve comparing survival rates between an experimental drug and placebo.

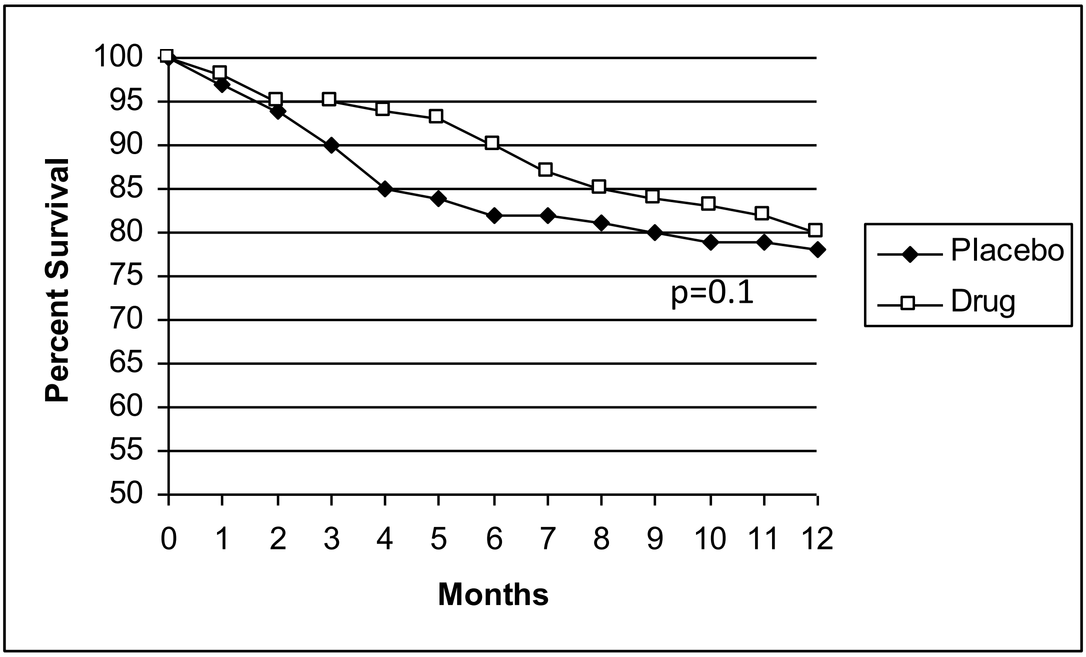

Based on the above Kaplan-Meier curve, what conclusion can you make about the drug?

A. The drug does not decrease mortality more than the placebo because the curves eventually converge.
B. The drug does not decrease mortality more than the placebo because p >0.05.
C. The drug does decrease mortality more than the placebo, but only between 2 and 12 months.
**D. None of the above.**

18. The odds ratio of developing an abdominal aortic aneurysm for a male smoker compared to his non-smoking counterpart is 3. If the probability of a non-smoker developing an abdominal aortic aneurysm is 10%, what is the probability that a smoker will develop an abdominal aortic aneurysm?

A. 20%
**B. 25%**
C. 30%
D. Cannot be calculated from the information given

*Lederle et al. The aneurysm detection and management study screening program: validation cohort and final results. Aneurysm Detection and Management Veterans Affairs Cooperative Study Investigators. Arch Intern Med. 2000;160(10):1425.*

### SPLIT TEST

**Six progressive levels in testing of evidence-based medicine (SPLIT) instrument (1 point for each answer)**

**EB-KN. To increase study power, one should increase:**

- **A. Sample size**
- B. P value
- C. Statistical test power
- D. Internal validity of research
- E. Specificity

**IV-UN. Why isn't arithmetic mean a representative measure of central tendency in a skewed distribution?**

- A. Because a skewed distribution does not have a measure of central tendency.
- B. Because it is difficult to present graphically.
- C. Because skewed distribution cannot be mathematically defined.
- **D. Because a single extreme result can significantly alter the mean.**
- E. Because calculating arithmetic mean includes squaring, which in case of skewed distribution disproportionally increases extreme values.

**CI.AN. What is the efficacy of treatment whose NNT is 5 (95% confidence interval 3-43)?**

- A. Small, because NNT is small and confidence interval includes 5.
- B. Large, because NNT is small and confidence interval includes 5.
- **C. Small, because NNT is small and confidence interval is large.**
- D. Large, because both NNT and confidence interval are large.
- E. Small, because NNT is large and confidence interval is small.

**Read summaries of the next two studies carefully and answer the questions EB-EV and IV-EV.**

**Study 1**

Association between daydreaming while driving and risk of a traffic accident in which driver holds the cilpability has been studied. A case-control study was conducted, including 955 drivers who were hurt in a traffic accident. Intensive daydreaming was found to be related to the liability for the traffic accident (17% in accidents in which drivers considered themselves responsible, and 9% in accidents where drivers thought they were not directly responsible), OR=2.12, 95% CI 1.37-3.28).

**Study 2**

The goal of the study was to investigate the relation between migraine and sudden cardiovascular events in women. A cohort of women was followed prospectively from 1989 to 2011. 90% of 115.541 initially included participants finished the study. Outcome was a „big "cardiovascular event: heart attack (myocardial infarction), a stroke (cerebrovascular insult or accident) or some other form of lethal cardiovascular morbidity. Migraine was related both to the increased risk of a big cardiovascular event (relative risk 1.50, 95% CI 1.33-1.69) and increased risk of death by cardiovascular morbidity (relative risk 1.79, 95% CI 1.33-2.32). Study results show there is a consistent relationship between migraine and cardiovascular disease.

**EB-EV. Mark the correct statement of the following:**

A. Study 1 has larger power and is more applicable.
B. Study 2 has larger power, but is less applicable.
**C. Study 1 has more confounding factors.**
D. Study 2 has smaller power and smaller applicability.
E. Both studies have equal power and applicability.

**IV-EV. Mark the correct statement of the following:**

A. **Study 2 had larger power because of the larger sample size.**
B. Study 2 shows that migraines have an influence on sudden cardiovascular events.
C. Study 1 was prospective.
D. Study 1 shows daydreaming causes traffic accidents.
E. Confidence interval of both studies are too wide.

**SD-EV. Why does a randomised controlled trial (RCT) provide better quality of evidence than a cohort study?**

A. RCT has a control group, and a cohort study does not.
B. RCT has balanced group sizes, and a cohort study does not.
C. RCT is cheaper than a cohort study.
D. RCT is shorter in duration than a cohort study.
**E. RCT controls exposure to investigated factor, and a cohort study does not.**

**IS-KN. The guideline for reporting cohort study results is called:**

A. COCHRANE.
B. PRISMA.
C. STARD.
D. CONSORT.
E. **STROBE.**

**IS-UN. You want to search databases for information on breast cancer. Searching the key word „breast“ retrieved 4200 articles, and key word „cancer“ retrieved 34150 articles. Which of the following statements is correct:**

A. Using those two key words and the operator AND will retrieve 38150 articles.
B. **By using the operator OR to combine „breast“ and „cancer“ will retrieve at least 4200 articles.**
C. Search strategy „breast NOT cancer“ will retrieve more than 4200 articles.
D. Search strategy „cancer NOT breast“ will retrieve 34150 articles.
E. By using the operator OR you will get fewer than 4200 articles.

**DS-EV. It is better to have high sensitivity and low specificity of a diagnostic test, than a low sensitivity and high specificity, if:**

A. The test is very cheap.
B. **The disease is very dangerous.**
C. Additional diagnostic procedures are very aggressive.
D. The disease is very rare.
E. The test is time consuming.

**Choose the correct combination of answers that fits the research plan. Each answer is made up of six parts of research plan: hypothesis, study design, inclusion criteria, exclusion criteria, main outcome measure and one potentially confounding factor.**

**S1. You want to know if eating ice cream too quickly causes headache.**

| Answer | Hypothesis | Type of study design | Exclusion criteria | Outcome measure | Confounding factor |
|---|---|---|---|---|---|
| A | Eating ice cream too quickly causes headaches. | Cross-over trial | Subjects with frequent migraines | Incidence of headache | Headache localization |
| B | Eating ice cream too quickly causes headaches. | Qualitative study | Subjects with frequent migraines | Incidence of headache | Headache localization |
| **C** | **Eating ice cream too quickly causes headaches.** | **Cross-over trial** | **Subjects with frequent migraines** | **Incidence of headache** | **Speed of eating ice cream** |
| D | Eating ice cream too quickly causes headaches. | Qualitative study | Subjects with frequent migraines | Incidence of headache | Speed of eating ice cream |
| E | Eating ice cream too quickly causes headaches. | Cross-over trial | Subjects with frequent migraines | Incidence of headache | Headache localization |

**IV-KN. Prevalence of disease in a population is best determined by:**
A. A randomized controlled trial (RCT).
B. A cohort study.
**C. A cross-sectional study.**
D. A qualitative study.
E. A retrospective study.

**IV-AN. Among given studies, choose one with design that is at the top of the pyramid of clinical relevance:**

A. To investigate attitudes towards mental health patients, the researchers conducted a phone survey on a sample of Croatian citizens, choosing the participants randomly from telephone directory.
B. In order to closely explore the relation between zinc intake and prostate cancer, the researchers compared a group of prostate cancer patients with a group of urologic patients without cancer and assessed differences in their zinc levels.
C. **To investigate which therapies help in alopecia areata treatment, the researchers analysed 17 randomized controlled trials of alopecia treatment.**
D. To check the efficacy of new alcohol rehabilitation treatment, participants were randomized into two groups, one that was treated with a new anxiety medication and a second group that that was given older and well known anxiety medication. Main outcome measure was relapse, i.e. dropping out of rehabilitation programme.
E. To determine if lower cardiovascular fitness is associated with sudden depression, 12 prospective cohort studies were analysed.

**SD-KN. Randomized controlled trial (RCT) outcome measures are all of the following except:**

A. Incidence.
B. **Prevalence.**
C. Event rate.
D. NNT.
E. Absolute risk.

**EB-AP. Prevalence of headache in one year on a sample of Split-Dalmatia County citizens was measured, and these are the results:**

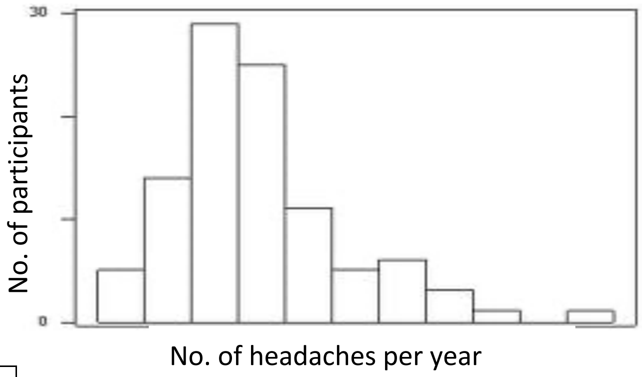

| | |
|---|---|
| Arithmetic mean | 16.8 |
| Median | 12 |
| Standard deviation | 2.79 |
| Interquartile range | 6 |
| Total range | 2-35 |

**Which measures should be used to describe the sample?**

A. Arithmetic mean and standard deviation (M=16.8 i SD=2.79).
B. Median and interquartile range (Md=12 i IQR=6).
C. Arithmetic mean and total range (M=16.8 and total range 2-35).
D. **All of 5 listed measures, depending on statistical test being used.**
E. Median and standard deviation (Md=12 i SD=2.79).

**IS-AN. Which part of a scientific article does the next sentence belong to: „Number of prepared questionnaires was defined according to hospital employee list (total of 750 physicians)“:**

A. Introduction.
B. **Methods.**
C. Results.
D. Discussion.
E. Appendix.

**DS-KN. The proportion of true negative results of a diagnostic test in the total number of healthy participants is called:**

A. Positive predictive value.
B. Precision.
C. Negative predictive value.
D. **Specificity.**
E. Sensitivity.

**IS-EV. Two systematic reviews were produced. The first one was conducted in 2009, and included 43 randomized controlled trials that tested efficacy of cognitive behavioural therapy in treating anxiety attacks. An update was not conducted. The second SR was performed in 2016, and included 10 RCTs that investigated influence of working with computers and eyesight worsening. Which of the following statements is correct:**

A. **To assess the reliability of the studies, it is necessary to look at their conclusions.**
B. First systematic review is more reliable because it includes RCTs.
C. Second systematic review is more reliable because it is more recent.
D. First study is more reliable because it includes more studies.
E. First study is more reliable because it includes more studies and more randomized controlled trials.

**CI-AP. A new treatment for social anxiety was investigated and it provided normal everyday functioning in 20% of the patients, compared to 10% of the patients who were treated by standard treatment. How many patients should be included in the new program to assure that one of them achieves normal everyday functioning?**

A. 2.
**B. 5.**
C. 20.
D. 10.
E. 15.

**S3. Choose a research plan that would provide you with answers on weekly alcohol consumption and its long-term consequences.**

| Answer | Goal of the study | Study design | Exclusion criteria | Outcome measure | Confounding factor |
|---|---|---|---|---|---|
| A | **To investigate long term consequences of alcohol consumption on a weekly basis.** | **Randomized controlled trial** | **Subjects who drink alcohol several times a week.** | **Incidence of subjects with side-effects** | **Subjects who drink different quantities of alcoholic beverages.** |
| B | To investigate long term consequences of alcohol consumption on a weekly basis. | Cohort study | Subjects who don't drink alcohol. | Incidence of subjects with side-effects | Subjects who drink various alcoholic beverages. |
| C | To investigate long term consequences of alcohol consumption on a weekly basis. | Case-control study | Subjects who drink various alcoholic beverages. | Odds ratio that measures association between weekly consumption of alcohol and its long term consequences | Subjects who drink different quantities of alcoholic beverages. |
| D | To investigate long term consequences of alcohol consumption on a weekly basis. | Cohort study | Subjects who drink alcohol several times a week. | Incidence of subjects with side-effects | Subjects who drink different quantities of alcoholic beverages. |
| E | There are long term consequences of alcohol consumption on a weekly basis. | Cross-sectional study | Subjects who drink various alcoholic beverages. | Prevalence of subjects with side-effects | Subjects who drink various alcoholic beverages. |

**CI-UN. A study of relation between consumption of black tea and weight loss in overweight participants gave the following results: group that consumed black tea for six months lost an average of 328 g (95% CI 125 – 922 g), and the group that did not lost an average of 121 g (95% CI 50 – 211 g). What conclusions can you draw from this research?**

A. Black tea helped in reducing bodyweight for an average of 207 g.

B. Black tea helped in reducing bodyweight, but the result is not statistically significant.
C. Black tea helped in reducing bodyweight and the result is statistically significant.
D. **We cannot make conclusions about efficacy of black tea from available data.**
E. Black tea did not help in reducing bodyweight.

**S5. You want to investigate the efficacy of corticosteroids in the treatment of chronic foot dermatitis in comparison to antibiotics, which are considered to be the golden standard.**

| **Answer** | **Study design** | **Inclusion criteria** | **Exclusion criteria** | **Outcome measure** | **Confounding factor** |
|---|---|---|---|---|---|
| A | RCT | Subjects with chronic foot dermatitis | Subjects who have psoriasis | NNT for corticosteroids | Subjects using both treatments |
| **B** | **Cohort study** | **Subjects with chronic foot dermatitis** | **Subjects who have a fungal infection** | **Relative risk reduction** | **Time which passes and aids treatment** |
| C | Crossover trial | Subjects who agreed to participate in the research | Subjects who have a fungal infection | NNT for corticosteroids | Effects of the first treatment to the current one |
| D | Systematic review of RCTs | Subjects with dermatitis | Subjects who have a fungal infection | Comparison of NNT for corticosteroids and for antibiotics | Possibility of a very few studies available |
| E | Systematic review of cohort studies | Subjects who do not have foot dermatitis | Subjects who have psoriasis | Absolute risk | Lack of a control group |

**SD-UN. While investigating a possible connection between marijuana smoking and lung cancer, it was found that the number of participants who smoked marijuana was significantly higher in the group who had lung cancer than the healthy control group (OR=1.3, 95% CI 1.1-1.7). Correct interpretation of this result is:**

A. Subjects who smoke marijuana are 10-70% less likely to get lung cancer.
B. **Subjects who smoke marijuana are 30% more likely to get lung cancer.**
C. Subjects who smoke marijuana are 30% less likely to get lung cancer.
D. There is no association between marijuana smoking and lung cancer.
E. No conclusion can be made on the association between marijuana smoking and lung cancer.

**EB-UN. While comparing the efficacy of drug A and drug B, it was found that drug B is significantly more efficient than drug A (P=0.017). If the study power was 0.8, what can you conclude from that information?**

A. That alpha value is 0.8.
B. That the sample is made of 4/5 of the population.
C. That the study includes 80% of the participants.
D. That the difference found between groups is significant at 80% probability level.
E. **That there is 80% probability of finding a difference between groups.**

**SD-AP. A conclusion that participants exposed to a factor A had lower incidence of disease was made out of:**

A. A cross-sectional study.
**B. A cohort study.**
C. A diagnostic study.
D. A physiological study.
E. A case-control study.

**DS-UN. Likelihood ratio for a test result was 12.4. We can conclude that:**

A. The subject has disease.
B. The subject does not have disease.
C. The test is not sensitive.
**D. The test has high specificity.**
E. The test finds disease in every 12th subject.

**CI-KN. The result of a case-control study is expressed as:**

**A. Odds ratio.**
B. Prevalence.
C. Absolute risk reduction.
D. Relative risk.
E. Positive predictive value.

**S6. Diagnostic test for colon cancer is very expensive and you want to develop a new one that would cost less. Parts of your research will be as follows:**

| Answer | Hypothesis | Study design | Inclusion criteria | Exclusion criteria | Outcome measure | Possible bias |
|---|---|---|---|---|---|---|
| **A.** | **There will be no difference between tests in the proportion of true results** | **Cross-sectional study** | **Subjects who want to find out if they have colon cancer** | **None** | **Specificity and sensitivity of the new test** | **Not all subjects are tested with the golden standard** |
| B. | Is there a difference between tests in the proportion of true results? | Cohort study | Subjects who have colon cancer | Subjects who do not have colon cancer | Number of subjects who develop colon cancer | Big attrition rate |
| C. | Is there a difference between tests in the proportion of true results? | RCT | Subjects who have colon cancer | Subjects who do not have colon cancer | Number of subjects who do not develop colon cancer | Small sample size |
| D. | There will be no difference between tests in the proportion of true results | Cohort study | Subjects who do not have colon cancer | Subjects who have colon cancer | Number of subjects who do not develop colon cancer | Not all subjects are tested with the golden standard |
| E. | New test will be better at diagnosis compared to the golden standard | Cross-sectional study | Subjects who apply for research | Subjects who have colon cancer | Specificity and sensitivity | Difficult to randomize |

**S2. Your patient wants to know how likely it is that he has prostate cancer. The patient does not suffer from any medical condition. You search PubMed to find research that would provide you with an answer to that question. What combination of search terms would you use?**

| Answer | Goal of the study | Study design | Exclusion criteria | Outcome measure | Confounding factor |
|---|---|---|---|---|---|
| **A** | **Patient does not have prostate cancer.** | **Cross-sectional study** | **Subjects who have several types of cancer.** | **Incidence of subjects with prostate cancer** | **Country of research** |
| B | Patient has prostate cancer. | Systematic review of RCTs | Subjects in good health. | Incidence of subjects with prostate cancer | Country of research |
| C | To check the likelihood of your patient not having prostate cancer. | Systematic review of cross-sectional studies | Subjects who have prostate cancer. | Prevalence of subjects with prostate cancer | Sample size |
| D | To check the likelihood of your patient having prostate cancer. | Systematic review of cohort studies | Subjects who have several types of cancer. | Incidence of subjects with prostate cancer | Sample size |
| E | To check the likelihood of your patient having prostate cancer. | Cross-sectional study | Subjects who have several types of cancer. | Prevalence of subjects with prostate cancer | Country of research |

**IV-AP. A physician wants to investigate differences in rates and numbers of metastasis in all of the hospital lung cancer patients, depending on their cancer stage (I.-IV.). The sample in such research will be:**

A. **Convenience sample.**
B. A random sample.
C. A systematic sample.
D. A stratified sample.
E. A cluster sample.

**SS-AN. In cohort studies, what is the advantage of prospective compared to historical prospective study design?**

A. A prospective study is cheaper.
B. In a prospective study participants can be randomized.
C. A prospective study has the possibility of analysing adverse events, and a historical study does not.
D. A prospective study requires smaller sample size.
E. **In a prospective study you can make the choice of most relevant and up to date outcome measures.**

**DS-AN. Among given values of the areas under the curve (AUC), choose the one that would fit the test that distinguishes between healthy and ill participants the least:**

A. 1.0
B. 0.7
**C. 0.5**
D. 0.3
E. 0.0

**S4. You want to see what is the probability of getting colon cancer if you sit more than 10 hours a day for a long period of time.**

| Answer | Hypothesis | Study design | Inclusion criteria | Exclusion criteria | Outcome measure |
|---|---|---|---|---|---|
| A | What is the probability of getting colon cancer if you spend more than 10 hours daily sitting for a longer periods of time? | Systematic review of randomized controlled trials | Subjects who do not have colon cancer | Subjects with advanced stages of colon cancer | Colon cancer diagnosis |
| **B** | **What is the probability of getting colon cancer if you spend more than 10 hours daily sitting for a long period of time?** | **Cohort study** | **Subjects who do not have colon cancer** | **Subjects who pass away during research** | **Incidence of colon cancer** |
| C | What is the probability of getting colon cancer if you spend more than 10 hours daily sitting for a long period of time? | Randomized controlled trial | Subjects who have colon cancer | Subjects who do not want to participate | NNT (for sitting) |
| D | What is the probability of getting colon cancer if you spend more than 10 hours daily sitting for a long period of time? | Cross-sectional study | Subjects with and without colon cancer | Subjects who do not spend more than 10 hours a day sitting | Prevalence |
| E | Subjects who spend more than 10 hours daily sitting for a long period of time are more likely to get colon cancer | Case-control study | Subjects with and without colon cancer | Smokers | Odds ratio for getting colon cancer |

**CI-EV. Which of the following findings has the highest clinical value?**

A. Lowering blood cholesterol levels from 8 mmol/L to 6 mmol/L; P=0.049.
**B. Lowering five-year mortality rate from prostate cancer for 7%; P=0.114.**
C. Lowering mean arterial pressure from 111 mmHg to 107 mmHg; P<0.001.
D. Lowering five-year mortality rate from 5th year breast cancer for 2%; P=0.037.
E. Lowering blood cholesterol levels from 6 mmol/L to 4.2 mmol/L; P=0.052.

**DS-AP. The prevalence of a disease is 10%. The diagnostic test used for it has 80% specificity and 90% sensitivity. If you get a positive result, what is the likelihood of actually having the disease?**

**A. Around 33%**
B. Around 30%
C. Around 38%
D. Around 90%
E. Around 80%

**EB-AN. Complete the summary with a conclusion.**

*Introduction:* To investigate if acute fear can actually „freeze the blood in your veins", researchers recruited 24 healthy volunteers (under 30 years).
*Methods:* Via random selection, 14 of the volunteers were sorted into a group that watched a horror movie before watching a documentary, and the remaining 10 volunteers watched the movies in reverse order. The movies were about 90 minutes long each and the volunteers watched them 7 days apart. Primary outcome was blood levels of coagulation factor VIII. Secondary outcome was participants' own experience of fear while watching the movies.
*Results:* Results show that, after watching the horror movie, mean difference in perceived fear was 5.4 units (95% CI 4.7-6.1) on a visual analogue scale (VAS), and blood levels of coagulation factor VII were significantly increased (mean difference 11.1 IU/dL (111 IU/L), 95% CI 1.2-21.0 IU/dL).

***Conclusion* is the following:**

A. There was a statistically significant difference in levels of coagulation factor VIII after watching the documentary.
**B. We cannot conclude with certainty, but the results indicate that horror movies increase blood levels of coagulation factor VII.**
C. There is not enough evidence to conclude that horror movies affect blood levels of coagulation factor VIII.
D. Fear can actually „freeze the blood in your veins".
E. There was a statistically significant increase of coagulation factor, but not of perceived fear.

**IS-AP. You want to find the article that described the study mentioned above. What combination of key words would you choose to find the exact same article?**

A. Healthy volunteers, documentary/horror movie, factor VIII.
B. Healthy volunteers, factor VIII, visual analogue scale, horror movie.
C. Healthy volunteers, horror movie, visual analogue scale, factor VIII.
**D. Healthy volunteers, horror movie, documentary, factor VIII.**
E. Horror movie, documentary, visual analogue scale, factor VIII.
