## Supplementary material for "Prospective study on the organization and efficiency of online journal club": in Supplementary 2

### Supplementary table: Detailed description of each session.

| № meeting | Date | Participants at the meeting | Members JC | Percent of visits, % | Lecture | Research | Format | Time (h:mm) | Specialty |
|---|---|---|---|---|---|---|---|---|---|
| 1 | 04.11.2020 | 15 | 19 | 79% | Evidence-based medicine | | Lecture | 2:00 | Evidence-based medicine |
| 2 | 14.11.2020 | 10 | 19 | 53% | basic definitions of statistics for reading articles | Wallentin L, Becker RC, Budaj A, Cannon CP, Emanuelsson H, Held C, Horrow J, Husted S, James S, Katus H, Mahaffey KW, Scirica BM, Skene A, Steg PG, Storey RF, Harrington RA; PLATO Investigators, Freij A, Thorsén M. Ticagrelor versus clopidogrel in patients with acute coronary syndromes. N Engl J Med. 2009 Sep 10;361(11):1045-57. doi: 10.1056/NEJMoa0904327. Epub 2009 Aug 30. PMID: 19717846. | Lecture, Discussion | 2:10 | Cardiology |
| 3 | 30.11.2020 | 8 | 19 | 42% | Descriptive statistics | Katelaris PH, Forbes GM, Talley NJ, Crotty B. A randomized comparison of quadruple and triple therapies for Helicobacter pylori eradication: The QUADRATE Study. Gastroenterology. 2002 Dec;123(6):1763-9. doi: 10.1053/gast.2002.37051. PMID: 12454831. | Lecture, Discussion | 2:18 | Gastroenterology |
| 4 | 20.12.2020 | 10 | 19 | 53% | Sampling calculation methods | Nordholm-Carstensen A, Schnack Rasmussen M, Krarup PM. Increased Leak Rates Following Stapled Versus Handsewn Ileocolic Anastomosis in Patients with Right-Sided Colon Cancer: A Nationwide Cohort Study. Dis Colon Rectum. 2019 May;62(5):542-548. doi: 10.1097/DCR.0000000000001289. PMID: 30489322. | Lecture, Discussion | 1:58 | Surgery |
| 5 | 24.12.2020 | 7 | 16 | 44% | Phases of clinical trials | Yeh RW, Valsdottir LR, Yeh MW, Shen C, Kramer DB, Strom JB, Secemsky EA, Healy JL, Domeier RM, Kazi DS, Nallamothu BK; PARACHUTE Investigators. Parachute use to prevent death and major trauma when jumping from aircraft: randomized controlled trial. BMJ. 2018 Dec 13;363:k5094. doi: 10.1136/bmj.k5094. Erratum in: BMJ. 2018 Dec 18;363:k5343. PMID: 30545967; PMCID: PMC6298200. | Lecture, Discussion | 1:57 | Christmas issue |

| № meeting | Date | Participants at the meeting | Members JC | Percent of visits, % | Lecture | Research | Format | Time (h:mm) | Specialty |
|---|---|---|---|---|---|---|---|---|---|
| 6 | 20.01.2021 | 6 | 14 | 43% | Non-inferiority trial | von Dach E, Albrich WC, Brunel AS, Prendki V, Cuvelier C, Flury D, Gayet-Ageron A, Huttner B, Kohler P, Lemmenmeier E, McCallin S, Rossel A, Harbarth S, Kaiser L, Bochud PY, Huttner A. Effect of C-Reactive Protein-Guided Antibiotic Treatment Duration, 7-Day Treatment, or 14-Day Treatment on 30-Day Clinical Failure Rate in Patients With Uncomplicated Gram-Negative Bacteremia: A Randomized Clinical Trial. JAMA. 2020 Jun 2;323(21):2160-2169. doi: 10.1001/jama.2020.6348. PMID: 32484534; PMCID: PMC7267846. | Lecture, Report (member JC) | 2:30 | Infection |
| 7 | 03.02.2021 | 11 | 12 | 92% | | HALT-IT Trial Collaborators. Effects of a high-dose 24-h infusion of tranexamic acid on death and thromboembolic events in patients with acute gastrointestinal bleeding (HALT-IT): an international randomised, double-blind, placebo-controlled trial. Lancet. 2020 Jun 20;395(10241):1927-1936. doi: 10.1016/S0140-6736(20)30848-5. PMID: 32563378; PMCID: PMC7306161. | Report (member JC) | 2:56 | Emergency medicine |
| 8 | 16.02.2021 | 9 | 12 | 75% | | Hague AL, Carr MP. Efficacy of an automated flossing device in different regions of the mouth. J Periodontol. 2007 Aug;78(8):1529-37. doi: 10.1902/jop.2007.060512. PMID: 17668972. | Discussion | 2:14 | Dentistry |
| 9 | 09.03.2021 | 27 | 33 | 82% | | Sauer R, Liersch T, Merkel S, Fietkau R, Hohenberger W, Hess C, Becker H, Raab HR, Villanueva MT, Witzigmann H, Wittekind C, Beissbarth T, Rödel C. Preoperative versus postoperative chemoradiotherapy for locally advanced rectal cancer: results of the German CAO/ARO/AIO-94 randomized phase III trial after a median follow-up of 11 years. J Clin Oncol. 2012 Jun 1;30(16):1926-33. doi: 10.1200/JCO.2011.40.1836. Epub 2012 Apr 23. PMID: 22529255. | Discussion | 2:24 | Oncology |
| 10 | 18.03.2021 | 21 | 33 | 64% | | Tsugawa Y, Jena AB, Figueroa JF, Orav EJ, Blumenthal DM, Jha AK. Comparison of Hospital Mortality and Readmission Rates for Medicare Patients Treated by Male vs Female Physicians. JAMA Intern Med. 2017 Feb | Discussion | 2:06 | Sociology |

| № meeting | Date | Participants at the meeting | Members JC | Percent of visits, % | Lecture | Research | Format | Time (h:mm) | Specialty |
|---|---|---|---|---|---|---|---|---|---|
| | | | | | | 1;177(2):206-213. doi: 10.1001/jamainternmed.2016.7875. PMID: 27992617; PMCID: PMC5558155. | | | |
| 11 | 31.03.2021 | 19 | 33 | 58% | Medical ethics | Mizoue T, Yoshimura T, Tokui N, Hoshiyama Y, Yatsuya H, Sakata K, Kondo T, Kikuchi S, Toyoshima H, Hayakawa N, Tamakoshi A, Ohno Y, Fujino Y, Kaneko S; Japan Collaborative Cohort Study Group. Prospective study of screening for stomach cancer in Japan. Int J Cancer. 2003 Aug 10;106(1):103-7. doi: 10.1002/ijc.11183. PMID: 12794764. | Lecture, Discussion (member JC) | 2:02 | Oncological screening |
| 12 | 06.05.2021 | 15 | 33 | 45% | p-value, "Why most research are false" | Adler L, Duncan E, Angrist B, Hemdal P, Rotrosen J, Slotnick V. Effects of a specific beta 2-receptor blocker in neuroleptic-induced akathisia. Psychiatry Res. 1989 Jan;27(1):1-4. doi: 10.1016/0165-1781(89)90002-4. PMID: 2564208. | Lecture, Report (member JC) | 2:15 | Psychiatry |
| 13 | 28.05.2021 | 12 | 33 | 36% | Protocols for communication with the patient | Okoli C, Van de Velde N, Richman B, Allan B, Castellanos E, Young B, Brough G, Eremin A, Corbelli GM, Mc Britton M, Hardy WD, de Los Rios P. Undetectable equals untransmittable (U = U): awareness and associations with health outcomes among people living with HIV in 25 countries. Sex Transm Infect. 2021 Feb;97(1):18-26. doi: 10.1136/sextrans-2020-054551. Epub 2020 Jul 30. PMID: 32732335; PMCID: PMC7841488. | Lecture, Report (member JC) | 2:28 | Infection, Sociology |
| 14 | 15.06.2021 | 8 | 30 | 27% | | Daly RM, Petrass N, Bass S, Nowson CA. The skeletal benefits of calcium- and vitamin D3-fortified milk are sustained in older men after withdrawal of supplementation: an 18-mo follow-up study. Am J Clin Nutr. 2008 Mar;87(3):771-7. doi: 10.1093/ajcn/87.3.771. PMID: 18326617. | Discussion | 2:00 | Nutritionology |
| 15 | 08.07.2021 | 14 | 26 | 54% | | Kamstrup PR, Tybjaerg-Hansen A, Steffensen R, Nordestgaard BG. Genetically elevated lipoprotein(a) and increased risk of myocardial infarction. JAMA. 2009 Jun 10;301(22):2331-9. doi: 10.1001/jama.2009.801. PMID: 19509380. | Discussion | 2:00 | Genetics |

| № meeting | Date | Participants at the meeting | Members JC | Percent of visits, % | Lecture | Research | Format | Time (h:mm) | Specialty |
|---|---|---|---|---|---|---|---|---|---|
| 16 | 08.08.2021 | 10 | 25 | 40% | Why do you need a journal club, PICO | Lemkes JS, Janssens GN, van der Hoeven NW, van de Ven PM, Marques KMJ, Nap A, van Leeuwen MAH, Appelman YEA, Knaapen P, Verouden NJW, Allaart CP, Brinckman SL, Saraber CE, Plomp KJ, Timmer JR, Kedhi E, Hermanides RS, Meuwissen M, Schaap J, van der Weerdt AP, van Rossum AC, Nijveldt R, van Royen N. Timing of revascularization in patients with transient ST-segment elevation myocardial infarction: a randomized clinical trial. Eur Heart J. 2019 Jan 14;40(3):283-291. doi: 10.1093/eurheartj/ehy651. PMID: 30371767. | Lecture, Discussion (member JC) | 2:13 | Cardiology |
| 17 | 14.09.2021 | 9 | 25 | 36% | | Tilly H, Castaigne S, Bordessoule D, Casassus P, Le Prisé PY, Tertian G, Desablens B, Henry-Amar M, Degos L. Low-dose cytarabine versus intensive chemotherapy in the treatment of acute nonlymphocytic leukemia in the elderly. J Clin Oncol. 1990 Feb;8(2):272-9. doi: 10.1200/JCO.1990.8.2.272. PMID: 2299370. | Discussion | 2:00 | Haematology |
| 18 | 01.10.2021 | 11 | 15 | 73% | Descriptive statistics, type I and II errors, p-value | Pala L, Dicembrini I, Mannucci E. Continuous subcutaneous insulin infusion vs modern multiple injection regimens in type 1 diabetes: an updated meta-analysis of randomized clinical trials. Acta Diabetol. 2019 Sep;56(9):973-980. doi: 10.1007/s00592-019-01326-5. Epub 2019 Apr 3. PMID: 30945047. | Lecture, Discussion (member JC) | 2:20 | Endocrinology |
| 19 | 14.10.2021 | 19 | 31 | 61% | Odds ratio, risk ratio, confidence interval. Clinical case | Stein GE. Comparison of single-dose fosfomycin and a 7-day course of nitrofurantoin in female patients with uncomplicated urinary tract infection. Clin Ther. 1999 Nov;21(11):1864-72. doi: 10.1016/S0149-2918(00)86734-X. PMID: 10890258. | Lecture, Discussion (member JC) | 3:00 | Urology |
| 20 | 28.10.2021 | 18 | 28 | 64% | | Terada K, Muro S, Sato S, Ohara T, Haruna A, Marumo S, Kinose D, Ogawa E, Hoshino Y, Niimi A, Terada T, Mishima M. Impact of gastro-oesophageal reflux disease symptoms on COPD exacerbation. Thorax. 2008 Nov;63(11):951-5. doi: 10.1136/thx.2007.092858. Epub 2008 Jun 5. PMID: 18535116. | Discussion | 2:50 | Pulmonology |

| № meeting | Date | Participants at the meeting | Members JC | Percent of visits, % | Lecture | Research | Format | Time (h:mm) | Specialty |
|---|---|---|---|---|---|---|---|---|---|
| 21 | 07.11.2021 | 16 | 26 | 62% | Limitations of evidence-based medicine | Ayeh SK, Abbey EJ, Khalifa BAA, Nudotor RD, Osei AD, Chidambaram V, Osuji N, Khan S, Salia EL, Oduwole MO, Yusuf HE, Lasisi O, Nosakhare E, Karakousis PC. Statins use and COVID-19 outcomes in hospitalized patients. PLoS One. 2021 Sep 10;16(9):e0256899. doi: 10.1371/journal.pone.0256899. PMID: 34506533; PMCID: PMC8432819. | Lecture, Discussion | 1:53 | Infection |
| 22 | 19.11.2021 | 15 | 25 | 60% | | Xie Y, Bowe B, Li T, Xian H, Yan Y, Al-Aly Z. Risk of death among users of Proton Pump Inhibitors: a longitudinal observational cohort study of United States veterans. BMJ Open. 2017 Jul 4;7(6):e015735. doi: 10.1136/bmjopen-2016-015735. PMID: 28676480; PMCID: PMC5642790. | Discussion | 2:01 | Gastroenterology |
| 23 | 02.12.2021 | 17 | 23 | 74% | | Watts CG, McLoughlin K, Goumas C, van Kemenade CH, Aitken JF, Soyer HP, Fernandez Peñas P, Guitera P, Scolyer RA, Morton RL, Menzies SW, Caruana M, Kang YJ, Mann GJ, Chakera AH, Madronio CM, Armstrong BK, Thompson JF, Cust AE. Association Between Melanoma Detected During Routine Skin Checks and Mortality. JAMA Dermatol. 2021 Nov 3:e213884. doi: 10.1001/jamadermatol.2021.3884. Epub ahead of print. PMID: 34730781; PMCID: PMC8567188. | Discussion | 2:34 | Dermatology |
| 24 | 16.12.2021 | 17 | 22 | 77% | | Zimerman A, Worsham C, Woo J, Jena AB. The need for speed: observational study of physician driving behaviors. BMJ. 2019 Dec 18;367:l6354. doi: 10.1136/bmj.l6354. PMID: 31852682; PMCID: PMC7191945. | Discussion | 2:05 | Christmas issue |
| 25 | 17.01.2022 | 12 | 22 | 55% | basic definitions of statistics for reading articles | | Lecture | 1:02 | Statistics |
| 26 | 27.01.2022 | 14 | 22 | 64% | | Svendsen JH, Diederichsen SZ, Højberg S, Krieger DW, Graff C, Kronborg C, Olesen MS, Nielsen JB, Holst AG, Brandes A, Haugan KJ, Køber L. Implantable loop recorder detection of atrial fibrillation to prevent stroke (The LOOP Study): a randomised controlled trial. Lancet. 2021 Oct 23;398(10310):1507-1516. doi: | Discussion + guest (head of PHS program at ITMO) | 2:06 | Cardiology |

| № meeting | Date | Participants at the meeting | Members JC | Percent of visits, % | Lecture | Research | Format | Time (h:mm) | Specialty |
|---|---|---|---|---|---|---|---|---|---|
| | | | | | | 10.1016/S0140-6736(21)01698-6. Epub 2021 Aug 29. Erratum in: Lancet. 2021 Oct 23;398(10310):1486. PMID: 34469766 | | | |
| 27 | 11.02.2022 | 14 | 21 | 67% | | Nawas MT, Shah GL, Feldman DR, Ruiz JD, Robilotti EV, Aslam AA, Dundas M, Kamboj M, Barker JN, Cho C, Chung DJ, Dahi PB, Giralt SA, Gyurkocza B, Lahoud OB, Landau HJ, Lin RJ, Mailankody S, Palomba ML, Papadopoulos EB, Politikos I, Ponce DM, Sauter CS, Shaffer BC, Scordo M, van den Brink MRM, Perales MA, Tamari R. Cellular Therapy During COVID-19: Lessons Learned and Preparing for Subsequent Waves. Transplant Cell Ther. 2021 May;27(5):438.e1-438.e6. doi: 10.1016/j.jtct.2021.02.011. Epub 2021 Feb 14. PMID: 33728417; PMCID: PMC7952254. | Discussion + guest (infectiologist) | 1:49 | Infection |
| 28 | 18.02.2022 | 16 | 20 | 80% | Research plan (from idea to results) | | Lecture | 2:10 | Statistics |
| 29 | 27.02.2022 | 15 | 20 | 75% | | Le Tourneau C, Delord JP, Gonçalves A, Gavoille C, Dubot C, Isambert N, Campone M, Trédan O, Massiani MA, Mauborgne C, Armanet S, Servant N, Bièche I, Bernard V, Gentien D, Jezequel P, Attignon V, Boyault S, Vincent-Salomon A, Servois V, Sablin MP, Kamal M, Paoletti X; SHIVA investigators. Molecularly targeted therapy based on tumour molecular profiling versus conventional therapy for advanced cancer (SHIVA): a multicentre, open-label, proof-of-concept, randomised, controlled phase 2 trial. Lancet Oncol. 2015 Oct;16(13):1324-34. doi: 10.1016/S1470-2045(15)00188-6. Epub 2015 Sep 3. PMID: 26342236. | Discussion + guest (oncologist, head of HSO) | 1:16 | Oncology |

| № meeting | Date | Participants at the meeting | Members JC | Percent of visits, % | Lecture | Research | Format | Time (h:mm) | Specialty |
|---|---|---|---|---|---|---|---|---|---|
| 30 | 15.03.2022 | 11 | 20 | 55% | | Raita Y, Goto T, Faridi MK, Brown DFM, Camargo CA Jr, Hasegawa K. Emergency department triage prediction of clinical outcomes using machine learning models. Crit Care. 2019 Feb 22;23(1):64. doi: 10.1186/s13054-019-2351-7. PMID: 30795786; PMCID: PMC6387562. | Discussion | 1:59 | Emergency medicine |
| 31 | 01.04.2022 | 29 | 35 | 83% | Nutrition of oncology patients | | Discussion + guest (gastroenterologist) | 2:45 | Gastroenterology |
| 32 | 18.04.2022 | 22 | 34 | 65% | Limitations of scientific thinking, p-value | Santini F, Pinchera A, Marsili A, Ceccarini G, Castagna MG, Valeriano R, Giannetti M, Taddei D, Centoni R, Scartabelli G, Rago T, Mammoli C, Elisei R, Vitti P. Lean body mass is a major determinant of levothyroxine dosage in the treatment of thyroid diseases. J Clin Endocrinol Metab. 2005 Jan;90(1):124-7. doi: 10.1210/jc.2004-1306. Epub 2004 Oct 13. PMID: 15483074. | Lecture, Discussion | 2:33 | Endocrinology |
| 33 | 28.04.2022 | 23 | 33 | 70% | Sequencing | NICUSeq Study Group, Krantz ID, Medne L, Weatherly JM, Wild KT, Biswas S, Devkota B, Hartman T, Brunelli L, Fishler KP, Abdul-Rahman O, Euteneuer JC, Hoover D, Dimmock D, Cleary J, Farnaes L, Knight J, Schwarz AJ, Vargas-Shiraishi OM, Wigby K, Zadeh N, Shinawi M, Wambach JA, Baldridge D, Cole FS, Wegner DJ, Urraca N, Holtrop S, Mostafavi R, Mroczkowski HJ, Pivnick EK, Ward JC, Talati A, Brown CW, Belmont JW, Ortega JL, Robinson KD, Brocklehurst WT, Perry DL, Ajay SS, Hagelstrom RT, Bennett M, Rajan V, Taft RJ. Effect of Whole-Genome Sequencing on the Clinical Management of Acutely Ill Infants With Suspected Genetic Disease: A Randomized Clinical Trial. JAMA Pediatr. 2021 Dec 1;175(12):1218-1226. doi: 10.1001/jamapediatrics.2021.3496. Erratum in: JAMA Pediatr. 2021 Dec 1;175(12):1295. PMID: 34570182; PMCID: PMC8477301. | Lecture, Discussion (member JC) | 2:29 | Genetics, Paediatrics |

| № meeting | Date | Participants at the meeting | Members JC | Percent of visits, % | Lecture | Research | Format | Time (h:mm) | Specialty |
|---|---|---|---|---|---|---|---|---|---|
| 34 | 13.05.2022 | 19 | 32 | 59% | | André MPE, Carde P, Viviani S, Bellei M, Fortpied C, Hutchings M, Gianni AM, Brice P, Casasnovas O, Gobbi PG, Zinzani PL, Dupuis J, Iannitto E, Rambaldi A, Brière J, Clément-Filliatre L, Heczko M, Valagussa P, Douxfils J, Depaus J, Federico M, Mounier N. Long-term overall survival and toxicities of ABVD vs BEACOPP in advanced Hodgkin lymphoma: A pooled analysis of four randomized trials. Cancer Med. 2020 Sep;9(18):6565-6575. doi: 10.1002/cam4.3298. Epub 2020 Jul 25. PMID: 32710498; PMCID: PMC7520354. | Discussion | 2:01 | Haematology |
| 35 | 27.06.2022 | 20 | 31 | 65% | | Blokhin B.M., Shamsheva O.V., Chernaya N.L., Sitnikov I.G., Lazareva S.G., Balzerovich N.B., Perminova O.A., Zhiglinskaya O.V., Koshavtseva M.Yu. Results of a multicentre double-blind placebo-controlled randomized trial of the liquid form of Anaferon for children in the treatment of acute upper respiratory tract infections. RosVestnPerinatoliPediatr2019;64:(4): 105-113(inRuss). DOI: 10.21508/1027-4065- 2019-64-4-105-113 | Discussion | 2:17 | Homeopathy |
| 36 | 15.07.2022 | 21 | 29 | 72% | Basic definitions of causal inference, bias (classification, example) | Kendler KS, Aggen SH, Li Y, Lewis CM, Breen G, Boomsma DI, Bot M, Penninx BW, Flint J. The similarity of the structure of DSM-IV criteria for major depression in depressed women from China, the United States and Europe. Psychol Med. 2015 Jul;45(9):1945-54. doi: 10.1017/S0033291714003067. Epub 2015 Mar 17. PMID: 25781917; PMCID: PMC4446696. | Lecture, Discussion (member JC) | 2:31 | Psychiatry |
| 37 | 28.07.2022 | 22 | 27 | 81% | Read statistical results | | Lecture + guest (head of statistical program at Bioinformatics institute) | 2:15 | Statistics |
| 38 | 10.08.2022 | 15 | 27 | 56% | | Saldanha G, Khanna A, O'Riordan M, Bamford M. The Width of Invasion in Malignant Melanoma Is a Novel Prognostic Feature That Accounts for Outcome Better Than Breslow Thickness. Am J Surg Pathol. 2020 | Discussion + guest (pathologist) | 1:49 | Pathology |

| № meeting | Date | Participants at the meeting | Members JC | Percent of visits, % | Lecture | Research | Format | Time (h:mm) | Specialty |
|---|---|---|---|---|---|---|---|---|---|
| | | | | | | Nov;44(11):1522-1527. doi: 10.1097/PAS.0000000000001529. PMID: 3260416 | | | |
| 39 | 29.08.2022 | 21 | 26 | 81% | | Downward P, Rasciute S, Kumar H. The effect of health on social capital; a longitudinal observation study of the UK. BMC Public Health. 2020 Apr 7;20(1):466. doi: 10.1186/s12889-020-08577-w. PMID: 32264853; PMCID: PMC7137318. | Discussion | 1:40 | Organization |
| 40 | 14.09.2022 | 19 | 25 | 76% | | Wang JQ, Wu GR, Wang Z, Dai XP, Li XR. Long-term clinical outcomes of statin use for chronic heart failure: a meta-analysis of 15 prospective studies. Heart Lung Circ. 2014 Feb;23(2):105-13. doi: 10.1016/j.hlc.2013.07.012. Epub 2013 Aug 17. PMID: 23962886. | Discussion | 2:14 | Cardiology |
| 41 | 01.10.2022 | 15 | 25 | 60% | | EuroSurg Collaborative. Intraperitoneal drain placement and outcomes after elective colorectal surgery: international matched, prospective, cohort study. Br J Surg. 2022 May 16;109(6):520-529. doi: 10.1093/bjs/znac069. PMID: 35576382. | Discussion | 2:34 | Surgery, Oncology |
| 42 | 18.10.2022 | 19 | 25 | 76% | | Watanabe R, Doodnaught G, Proulx C, Auger JP, Monteiro B, Dumais Y, Beauchamp G, Segura M, Steagall P. A multidisciplinary study of pain in cats undergoing dental extractions: A prospective, blinded, clinical trial. PLoS One. 2019 Mar 1;14(3):e0213195. doi: 10.1371/journal.pone.0213195. PMID: 30822336; PMCID: PMC6396900. | Discussion | 2:18 | Veterinary |
| 43 | 02.11.2022 | 16 | 25 | 64% | | Kim KH, Kwon SH, Sim WY, Lew BL. The Study of Relationship between Anatomical Sites and Depth of the Lipoma. Ann Dermatol. 2021 Dec;33(6):562-567. doi: 10.5021/ad.2021.33.6.562. Epub 2021 Nov 4. PMID: 34858008; PMCID: PMC8577906. | Discussion | 1:29 | Dermatology |
| 44 | 16.11.2022 | 22 | 24 | 92% | DeSci, DAO | | Lecture + guest (head of Jocelyn DAO) | 2:02 | Organization |

| № meeting | Date | Participants at the meeting | Members JC | Percent of visits, % | Lecture | Research | Format | Time (h:mm) | Specialty |
|---|---|---|---|---|---|---|---|---|---|
| 45 | 07.12.2022 | 18 | 24 | 75% | | Morgan DE, Baron TH, Smith JK, Robbin ML, Kenney PJ. Pancreatic fluid collections prior to intervention: evaluation with MR imaging compared with CT and US. Radiology. 1997 Jun;203(3):773-8. doi: 10.1148/radiology.203.3.9169703. PMID: 9169703. | Discussion | 2:15 | Gastroenterology, Surgery |
| 46 | 22.12.2022 | 15 | 24 | 63% | | Martikainen P, Korhonen K, Tarkiainen L. Heavy metal toxicity and mortality-association between density of heavy metal bands and cause specific hospital admissions and mortality: population based cohort study. BMJ. 2021 Dec 15;375:e067633. doi: 10.1136/bmj-2021-067633. PMID: 34911746. | Discussion | 1:48 | Christmas issue |
| 47 | 18.01.2023 | 33 | 41 | 80% | | Short TG, Campbell D, Frampton C, Chan MTV, Myles PS, Corcoran TB, Sessler DI, Mills GH, Cata JP, Painter T, Byrne K, Han R, Chu MHM, McAllister DJ, Leslie K; Australian and New Zealand College of Anaesthetists Clinical Trials Network; Balanced Anaesthesia Study Group. Anaesthetic depth and complications after major surgery: an international, randomised controlled trial. Lancet. 2019 Nov 23;394(10212):1907-1914. doi: 10.1016/S0140-6736(19)32315-3. Epub 2019 Oct 20. PMID: 31645286. | Discussion | 2:35 | Anaesthesiology |
| 48 | 08.02.2023 | 31 | 41 | 76% | | Tilly H, Morschhauser F, Sehn LH, Friedberg JW, Trněný M, Sharman JP, Herbaux C, Burke JM, Matasar M, Rai S, Izutsu K, Mehta-Shah N, Oberic L, Chauchet A, Jurczak W, Song Y, Greil R, Mykhalska L, Bergua-Burgués JM, Cheung MC, Pinto A, Shin HJ, Hapgood G, Munhoz E, Abrisqueta P, Gau JP, Hirata J, Jiang Y, Yan M, Lee C, Flowers CR, Salles G. Polatuzumab Vedotin in Previously Untreated Diffuse Large B-Cell Lymphoma. N Engl J Med. 2022 Jan 27;386(4):351-363. doi: 10.1056/NEJMoa2115304. Epub 2021 Dec 14. PMID: 34904799. | Discussion (member JC) | 2:42 | Haematology |

| № meeting | Date | Participants at the meeting | Members JC | Percent of visits, % | Lecture | Research | Format | Time (h:mm) | Specialty |
|---|---|---|---|---|---|---|---|---|---|
| 49 | 22.02.2023 | 24 | 41 | 59% | Genetic methods | Arndt AK, Schafer S, Drenckhahn JD, Sabeh MK, Plovie ER, Caliebe A, Klopocki E, Musso G, Werdich AA, Kalwa H, Heinig M, Padera RF, Wassilew K, Bluhm J, Harnack C, Martitz J, Barton PJ, Greutmann M, Berger F, Hubner N, Siebert R, Kramer HH, Cook SA, MacRae CA, Klaassen S. Fine mapping of the 1p36 deletion syndrome identifies mutation of PRDM16 as a cause of cardiomyopathy. Am J Hum Genet. 2013 Jul 11;93(1):67-77. doi: 10.1016/j.ajhg.2013.05.015. Epub 2013 Jun 13. PMID: 23768516; PMCID: PMC3710750. | Lecture, Discussion (member JC) | 2:45 | Genetics |
| 50 | 09.03.2023 | 29 | 40 | 73% | | Staibano P, Forner D, Noel CW, Zhang H, Gupta M, Monteiro E, Sawka AM, Pasternak JD, Goldstein DP, de Almeida JR. Ultrasonography and Fine-Needle Aspiration in Indeterminate Thyroid Nodules: A Systematic Review of Diagnostic Test Accuracy. Laryngoscope. 2022 Jan;132(1):242-251. doi: 10.1002/lary.29778. Epub 2021 Aug 19. PMID: 34411290. | Discussion + guest (endocrinologist) | 2:08 | Endocrinology |
| 51 | 07.04.2023 | 25 | 36 | 69% | | Kristich CJ, Wells CL, Dunny GM. A eukaryotic-type Ser/Thr kinase in Enterococcus faecalis mediates antimicrobial resistance and intestinal persistence. Proc Natl Acad Sci U S A. 2007 Feb 27;104(9):3508-13. doi: 10.1073/pnas.0608742104. Epub 2007 Feb 20. PMID: 17360674; PMCID: PMC1805595. | Discussion | 1:40 | Infection |
| 52 | 28.04.2023 | 25 | 35 | 71% | | Cuijpers P, Reynolds CF 3rd, Donker T, Li J, Andersson G, Beekman A. Personalized treatment of adult depression: medication, psychotherapy, or both? A systematic review. Depress Anxiety. 2012 Oct;29(10):855-64. doi: 10.1002/da.21985. Epub 2012 Jul 19. PMID: 22815247. | Discussion | 2:21 | Psychiatry |
| 53 | 16.05.2023 | 21 | 33 | 64% | basic definitions of statistics for reading articles | | Lecture | 1:48 | Statistics |

| № meeting | Date | Participants at the meeting | Members JC | Percent of visits, % | Lecture | Research | Format | Time (h:mm) | Specialty |
|---|---|---|---|---|---|---|---|---|---|
| 54 | 01.06.2023 | 23 | 33 | 70% | | Huybrechts KF, Bateman BT, Pawar A, Bessette LG, Mogun H, Levin R, Li H, Motsko S, Scantamburlo Fernandes MF, Upadhyaya HP, Hernandez-Diaz S. Maternal and fetal outcomes following exposure to duloxetine in pregnancy: cohort study. BMJ. 2020 Feb 19;368:m237. doi: 10.1136/bmj.m237. PMID: 32075794; PMCID: PMC7190016. | Discussion | 2:33 | Obstetrics and gynecology |
| 55 | 03.07.2023 | 22 | 31 | 71% | | Adamson AS, Suarez EA, Welch HG. Estimating Overdiagnosis of Melanoma Using Trends Among Black and White Patients in the US. JAMA Dermatol. 2022 Apr 1;158(4):426-431. doi: 10.1001/jamadermatol.2022.0139. PMID: 35293957; PMCID: PMC8928089. | Discussion | 1:39 | Dermatology |
| 56 | 18.07.2023 | 20 | 31 | 65% | | Mandal R, Samstein RM, Lee KW, Havel JJ, Wang H, Krishna C, Sabio EY, Makarov V, Kuo F, Blecua P, Ramaswamy AT, Durham JN, Bartlett B, Ma X, Srivastava R, Middha S, Zehir A, Hechtman JF, Morris LG, Weinhold N, Riaz N, Le DT, Diaz LA Jr, Chan TA. Genetic diversity of tumors with mismatch repair deficiency influences anti-PD-1 immunotherapy response. Science. 2019 May 3;364(6439):485-491. doi: 10.1126/science.aau0447. PMID: 31048490; PMCID: PMC6685207. | Discussion | 2:19 | Pathology |
| 57 | 05.08.2023 | 17 | 30 | 57% | | Ilic D, de Voogt A, Oldroyd J. The use of journal clubs to teach evidence-based medicine to health professionals: A systematic review and meta-analysis. J Evid Based Med. 2020 Feb;13(1):42-56. doi: 10.1111/jebm.12370. Epub 2020 Jan 17. PMID: 31951092. | Discussion | 2:08 | Education |
| 58 | 06.10.2023 | 19 | 26 | 73% | | Whiteman DC, Olsen CM, MacGregor S, Law MH, Thompson B, Dusingize JC, Green AC, Neale RE, Pandeya N; QSkin Study. The effect of screening on melanoma incidence and biopsy rates. Br J Dermatol. 2022 Oct;187(4):515-522. doi: 10.1111/bjd.21649. Epub 2022 Jun 28. PMID: 35531668; PMCID: PMC9796145. | Discussion (member JC) | 2:21 | Diagnostics |

| № meeting | Date | Participants at the meeting | Members JC | Percent of visits, % | Lecture | Research | Format | Time (h:mm) | Specialty |
|---|---|---|---|---|---|---|---|---|---|
| 59 | 17.10.2023 | 15 | 26 | 58% | | Kearon C, de Wit K, Parpia S, Schulman S, Afilalo M, Hirsch A, Spencer FA, Sharma S, D'Aragon F, Deshaies JF, Le Gal G, Lazo-Langner A, Wu C, Rudd-Scott L, Bates SM, Julian JA; PEGeD Study Investigators. Diagnosis of Pulmonary Embolism with d-Dimer Adjusted to Clinical Probability. N Engl J Med. 2019 Nov 28;381(22):2125-2134. doi: 10.1056/NEJMoa1909159. PMID: 31774957. | Discussion | 2:20 | Emergency medicine |
| 60 | 03.11.2023 | 16 | 26 | 62% | | Zhang Y, Li L, Guo C, Mu D, Feng B, Zuo X, Li Y. Effects of probiotic type, dose and treatment duration on irritable bowel syndrome diagnosed by Rome III criteria: a meta-analysis. BMC Gastroenterol. 2016 Jun 13;16(1):62. doi: 10.1186/s12876-016-0470-z. PMID: 27296254; PMCID: PMC4907258. | Discussion | 2:03 | Gastroenterology |
| 61 | 12.12.2023 | 37 | 43 | 86% | Designs of clinical study | Poole JE, Bahnson TD, Monahan KH, Johnson G, Rostami H, Silverstein AP, Al-Khalidi HR, Rosenberg Y, Mark DB, Lee KL, Packer DL; CABANA Investigators and ECG Rhythm Core Lab. Recurrence of Atrial Fibrillation After Catheter Ablation or Antiarrhythmic Drug Therapy in the CABANA Trial. J Am Coll Cardiol. 2020 Jun 30;75(25):3105-3118. doi: 10.1016/j.jacc.2020.04.065. PMID: 32586583; PMCID: PMC8064404. | Lecture, Discussion (member JC) | 2:52 | Cardiology |
| 62 | 28.12.2023 | 28 | 43 | 67% | Basic definitions of causal inference, p-value, confidence interval | Klamer K. Analysis of Barbie medical and science career dolls: descriptive quantitative study. BMJ. 2023 Dec 18;383:e077276. doi: 10.1136/bmj-2023-077276. PMID: 38110233; PMCID: PMC10728597. | Lecture, Discussion | 2:03 | Christmas issue |
| 63 | 17.01.2024 | 33 | 43 | 77% | The position of the TP53 gene in the development of cancer | | Lecture + guest (oncopathomorphologist) | 2:28 | Pathology |

| № meeting | Date | Participants at the meeting | Members JC | Percent of visits, % | Lecture | Research | Format | Time (h:mm) | Specialty |
|---|---|---|---|---|---|---|---|---|---|
| 64 | 02.02.2024 | 27 | 42 | 64% | AI as a diagnostic method for lung cancer | Liu M, Wu J, Wang N, Zhang X, Bai Y, Guo J, Zhang L, Liu S, Tao K. The value of artificial intelligence in the diagnosis of lung cancer: A systematic review and meta-analysis. PLoS One. 2023 Mar 23;18(3):e0273445. doi: 10.1371/journal.pone.0273445. PMID: 36952523; PMCID: PMC10035910. | Lecture, Discussion (member JC) | 2:08 | Diagnostics |
| 65 | 01.03.2024 | 28 | 40 | 70% | | Svenson MRE, Freeman TP, Maynard OM. The Effect of Conflicting Public Health Guidance on Smokers' and Vapers' E-cigarette Harm Perceptions. Nicotine Tob Res. 2022 Nov 12;24(12):1945-1950. doi: 10.1093/ntr/ntac163. PMID: 35793536; PMCID: PMC9653072. | Discussion | 2:21 | Public health |
| 66 | 19.03.2024 | 28 | 40 | 70% | | Noah DL, Drenzek CL, Smith JS, Krebs JW, Orciari L, Shaddock J, Sanderlin D, Whitfield S, Fekadu M, Olson JG, Rupprecht CE, Childs JE. Epidemiology of human rabies in the United States, 1980 to 1996. Ann Intern Med. 1998 Jun 1;128(11):922-30. doi: 10.7326/0003-4819-128-11-199806010-00012. PMID: 9634432. | Discussion | 2:00 | Infection |
| 67 | 25.04.2024 | 26 | 39 | 67% | | Lai CY, Wong MKW, Tong WH, Chu SY, Lau KY, Tan AML, Hui LL, Lao TTH, Leung TY. Effectiveness of a childbirth massage programme for labour pain relief in nulliparous pregnant women at term: a randomised controlled trial. Hong Kong Med J. 2021 Dec;27(6):405-412. doi: 10.12809/hkmj208629. Epub 2021 Dec 17. PMID: 34924363. | Discussion | 2:35 | Obstetrics and Gynecology |
| 68 | 11.05.2024 | 27 | 38 | 71% | Landscape in data science | Rita González-Márquez, Luca Schmidt, Benjamin M. Schmidt, Philipp Berens, Dmitry Kobak. The landscape of biomedical research. Patterns. 2024. doi: 10.1016/j.patter.2024.100968. | Lecture, Discussion + guest (data analytics) | 2:31 | Data science |
| 69 | 06.06.2024 | 26 | 38 | 68% | | Sun J, Li Z, Liu S, Xia T, Shen J. Biodegradable magnesium screw, titanium screw and direct embedding fixation in pedicled vascularized iliac bone graft transfer for osteonecrosis of the femoral head: a randomized controlled study. J Orthop Surg Res. 2023 Jul 22;18(1):523. doi: 10.1186/s13018-023-04012-z. PMID: 37481538; PMCID: PMC10363316. | Discussion | 2:06 | Traumatology |

| № meeting | Date | Participants at the meeting | Members JC | Percent of visits, % | Lecture | Research | Format | Time (h:mm) | Specialty |
|---|---|---|---|---|---|---|---|---|---|
| 70 | 21.06.2024 | 27 | 36 | 75% | why should a doctor be able to read articles critically | | Lecture (member JC) | 2:19 | Statistics |
| 71 | 16.07.2024 | 27 | 35 | 77% | Association measures in statistics | | Lecture | 2:05 | Statistics |
| 72 | 15.08.2024 | 13 | 33 | 39% | | Wilding JPH, Batterham RL, Calanna S, Davies M, Van Gaal LF, Lingvay I, McGowan BM, Rosenstock J, Tran MTD, Wadden TA, Wharton S, Yokote K, Zeuthen N, Kushner RF; STEP 1 Study Group. Once-Weekly Semaglutide in Adults with Overweight or Obesity. N Engl J Med. 2021 Mar 18;384(11):989-1002. doi: 10.1056/NEJMoa2032183. Epub 2021 Feb 10. PMID: 33567185. | Discussion | 1:56 | Endocrinology |
| 73 | 10.09.2024 | 20 | 32 | 63% | | De La Fouchardière C, Malka D, Cropet C, Chabaud S, Raimbourg J, Botsen D, Launay S, Evesque L, Vienot A, Perrier H, Jary M, Rinaldi Y, Coutzac C, Bachet JB, Neuzillet C, Williet N, Desgrippes R, Grainville T, Aparicio T, Peytier A, Lecomte T, Roth GS, Thirot-Bidault A, Lachaux N, Bouché O, Ghiringhelli F. Gemcitabine and Paclitaxel Versus Gemcitabine Alone After 5-Fluorouracil, Oxaliplatin, and Irinotecan in Metastatic Pancreatic Adenocarcinoma: A Randomized Phase III PRODIGE 65-UCGI 36-GEMPAX UNICANCER Study. J Clin Oncol. 2024 Mar 20;42(9):1055-1066. doi: 10.1200/JCO.23.00795. Epub 2024 Jan 17. PMID: 38232341. | Discussion | 1:50 | Oncology |
| 74 | 08.10.2024 | 22 | 27 | 81% | | Niiranen TJ, Kalesan B, Hamburg NM, Benjamin EJ, Mitchell GF, Vasan RS. Relative Contributions of Arterial Stiffness and Hypertension to Cardiovascular Disease: The Framingham Heart Study. J Am Heart Assoc. 2016 Oct 26;5(11):e004271. doi: | Discussion | 2:23 | Epidemiology |

| № meeting | Date | Participants at the meeting | Members JC | Percent of visits, % | Lecture | Research | Format | Time (h:mm) | Specialty |
| --- | --- | --- | --- | --- | --- | --- | --- | --- | --- |
| | | | | | | 10.1161/JAHA.116.004271. PMID: 27912210; PMCID: PMC5210358. | | | |
| 75 | 24.10.2024 | 23 | 27 | 85% | basic definitions of statistics for reading articles | | Lecture + guest (biostatistics) | 1:50 | Statistics |
| 76 | 12.11.2024 | 20 | 27 | 74% | | Liu L, Xue Y, Chen Y, Chen T, Zhong J, Shao X, Chen J. Acne and risk of mental disorders: A two-sample Mendelian randomization study based on large genome-wide association data. Front Public Health. 2023 Mar 31;11:1156522. doi: 10.3389/fpubh.2023.1156522. PMID: 37064666; PMCID: PMC10102334. | Discussion | 1:47 | Dermatology |
| 77 | 28.11.2024 | 32 | 32 | 71% | Evidence-based medicine | | Lecture | 1:53 | Evidence-based medicine |
| 78 | 20.12.2024 | 35 | 35 | 65% | Causal inference (DAG) | | Lecture + guest (biostatistics) | 2:58 | Statistics |
| 79 | 27.12.2024 | 25 | 25 | 46% | | Flynn SG, Park RS, Jena AB, Staffa SJ, Kim SY, Clarke JD, Pham IV, Lukovits KE, Huang SX, Sideridis GD, Bernier RS, Fiadjoe JE, Weinstock PH, Peyton JM, Stein ML, Kovatsis PG. Coaching inexperienced clinicians before a high stakes medical procedure: randomized clinical trial. BMJ. 2024 Dec 16;387:e080924. doi: 10.1136/bmj-2024-080924. PMID: 39681397; PMCID: PMC11648086. | Discussion | 1:59 | Christmas issue |

| № meeting | Date | Participants at the meeting | Members JC | Percent of visits, % | Lecture | Research | Format | Time (h:mm) | Specialty |
|---|---|---|---|---|---|---|---|---|---|
| 80 | 23.01.2025 | 44 | 44 | 83% | Estimand, estimate, estimator | | Lecture | 2:17 | Statistics |
| 81 | 30.01.2025 | 34 | 34 | 64% | Distribution and confidence interval | | Lecture | 2:18 | Statistics |
| 82 | 06.03.2025 | 37 | 37 | 70% | Hypothesis testing | | Lecture | 2:28 | Statistics |
| 83 | 20.03.2025 | 29 | 29 | 56% | Type error I and II | | Lecture | 2:18 | Statistics |
| 84 | 08.04.2025 | 29 | 29 | 57% | | Munkholm K, Paludan-Müller AS, Boesen K. Considering the methodological limitations in the evidence base of antidepressants for depression: a reanalysis of a network meta-analysis. BMJ Open. 2019 Jun 27;9(6):e024886. doi: 10.1136/bmjopen-2018-024886. PMID: 31248914; PMCID: PMC6597641. | Discussion | 1:57 | Psychiatry |
| 85 | 15.05.2025 | 24 | 24 | 50% | | Van den Bossche J, Devreese K, Malfait R, Van de Vyvere M, Wauters A, Neeis H, De Schouwer P. Reference intervals for a complete blood count determined on different automated haematology analysers: Abx Pentra 120 Retic, Coulter Gen-S, Sysmex SE 9500, Abbott Cell Dyn 4000 and Bayer Advia 120. Clin Chem Lab Med. 2002 Jan;40(1):69-73. doi: 10.1515/CCLM.2002.014. PMID: 11916274. | Discussion | 2:09 | Diagnostics |

| № meeting | Date | Participants at the meeting | Members JC | Percent of visits, % | Lecture | Research | Format | Time (h:mm) | Specialty |
|---|---|---|---|---|---|---|---|---|---|
| 86 | 29.05.2025 | 24 | 24 | 53% | Pharmacoeconomics |  | Lecture (member JC) | 1:51 | Pharmacoeconomics |
| 87 | 09.06.2025 | 23 | 23 | 56% |  | Rosettie, Katherine L et al. "Cost-effectiveness of HPV vaccination in 195 countries: A meta-regression analysis." PloS one vol. 16,12 e0260808. 20 Dec. 2021, doi:10.1371/journal.pone.0260808 | Discussion | 1:52 | Pharmacoeconomics |
| 88 | 27.06;2025 | 28 | 28 | 74% | Hypothesis testing |  | Lecture + guest (biostatistics) | 02:35 | Statistics |
| 89 | 18.09.2025 | 25 | 25 | 69% | Causal inference |  | Lecture | 01:29 | Statistics |
| 90 | 14.10.2025 | 28 | 28 | 78% |  | Valtuille Z, Trebossen V, Ouldali N, et al. The hidden pandemic: changes in outpatient mental health care among French children and adolescents. Eur Child Adolesc Psychiatry. Published online September 8, 2025. doi:10.1007/s00787-025-02847-x | Discussion | 2:03 | Psychiatry |
| 91 | 08.11.2025 | 23 | 23 | 64% |  | Carolan A, Hynes-Ryan C, Agarwal SM, et al. Metformin for the Prevention of Antipsychotic-Induced Weight Gain: Guideline Development and Consensus Validation. Schizophr Bull. 2025;51(5):1193-1205. doi:10.1093/schbul/sbae205 | Discussion + guest (endocrinologist) | 1:50 | Endocrinology |
| 92 | 28.11.2025 | 17 | 17 | 49% |  | Siegel RL, Kratzer TB, Giaquinto AN, Sung H, Jemal A. Cancer statistics, 2025. CA Cancer J Clin. 2025;75(1):10-45. doi:10.3322/caac.21871 | Discussion | 2:27 | Epidemiology |

| № meeting | Date | Participants at the meeting | Members JC | Percent of visits, % | Lecture | Research | Format | Time (h:mm) | Specialty |
|---|---|---|---|---|---|---|---|---|---|
| 93 | 09.12.2025 | 14 | 14 | 41% | | Støer NC, Botteri E, Lindemann K, Langseth H, Fortner RT. Low-dose aspirin and non-aspirin non-steroidal anti-inflammatory drugs and epithelial ovarian cancer survival: a registry-based cohort study in Norway. BMC Cancer. 2025;25(1):807. Published 2025 Apr 30. doi:10.1186/s12885-025-14168-y | Discussion | 1:18 | Oncology |
| 94 | 29.12.2025 | 23 | 23 | 68% | | Díez-Vidal A, Arribas JR. How recent is recent? Retrospective analysis of suspiciously timeless citations. BMJ. 2025;391:e086941. Published 2025 Dec 11. doi:10.1136/bmj-2025-086941 | Discussion | 1:57 | Christmas issue |
| 95 | 05.02.2026 | 18 | 18 | 56% | | Hernández G, Paredes I, Moran F, et al. Effect of postextubation noninvasive ventilation with active humidification vs high-flow nasal cannula on reintubation in patients at very high risk for extubation failure: a randomized trial. Intensive Care Med. 2022;48(12):1751-1759. doi:10.1007/s00134-022-06919-3 | Discussion | 3:09 | Anaesthesiology |
| 96 | 21.02.2026 | 20 | 20 | 69% | | CURRENT-OASIS 7 Investigators, Mehta SR, Bassand JP, et al. Dose comparisons of clopidogrel and aspirin in acute coronary syndromes. N Engl J Med. 2010;363(10):930-942. doi:10.1056/NEJMoa0909475 | Discussion | 2:35 | Cardiology |
